# Discrete dynamic survival analysis of epidemic case line lists

**DOI:** 10.64898/2026.11.10.26367756

**Authors:** Sandra Montes-Olivas, Sam Abbott, Sebastian Funk, Michael B. Gravenor, Simon D.W. Frost

## Abstract

Case line lists, which record dates of onset, recovery, and hospitalisation, are among the most granular data in infectious disease epidemiology and can provide insights into transmission dynamics. Dynamic survival analysis (DSA) fits a deterministic, differential-equation-based model to line lists to estimate parameters such as infectivity and the infectious period. Infectious periods are usually not exponential, and incorporating this into mechanistic models for line-list data often requires partial differential equation (PDE) schemes that can be difficult to implement and must be customised for each model, which may limit their use. Here, we propose Discrete Dynamic Survival Analysis (DDSA), a discrete-time DSA for case line lists based on difference equations. It matches calendar-day data directly without interpolation, and its log-likelihood is differentiable via forward-mode automatic differentiation, enabling off-the-shelf gradient-based samplers without model-specific numerical infrastructure. Non-exponential infectious periods are accommodated through a boxcar model or a discrete analogue of the “device of stages”. We apply the DDSA method to three historical outbreaks of influenza A(H1N1), Ebola, and measles. DDSA estimates of transmission parameters were close to published analyses where these were available, an agreement that partly reflects shared modelling choices, and the method handles incomplete line lists with missing onset or recovery dates. Synthetic experiments show that recovery intervals identify the infectious period, while infection times and final size constrain *R*_0_ when the population at risk is known. Discrete DSA provides a simple, extensible framework for simulation and inference of infectious disease dynamics.

## 1 Introduction

Mathematical models can offer insights into the transmission of infectious disease, allowing us to estimate key epidemiological quantities such as the reproductive number, forecast the number of new cases [1], and explore counterfactual scenarios, such as the impact of school closures or vaccination campaigns[2, 3]. Many models use aggregated case data i.e., counts of new cases per day or week, to infer model parameters. However, deeper insights can be gained through the use of case line lists, which record individual-level event times such as symptom onset, hospitalisation, recovery, or death [4, 5]. While aggregate counts reveal general trends, line lists provide detailed tracking of individual events, allowing the estimation of key time dynamics, such as the infectious period.

In current practice, line-list data are analysed in several ways: direct survival analysis with parametric (e.g., exponential, lognormal or Weibull) natural-history interval models, which lack a mechanistic link to population-level transmission dynamics [6, 7]. Individual-based simulation models, which can capture fine-scale contact structure but are computationally intensive and difficult to calibrate via gradient-based methods [8, 9], and aggregation of the line list to daily counts, discarding individual-level timing information [10, 11]. Each method presents trade-offs in realism, computational feasibility, and data accuracy.

KhudaBukhsh et al. [12] proposed a modelling paradigm, originally introduced as Survival Dynamical Systems (SDS) but now known as Dynamic Survival Analysis (DSA) [13–15], to fit dynamical models of disease transmission to line lists using concepts borrowed from survival analysis. By assuming a large population size, DSA simplifies the analysis of line lists by using a mean-field model described by differential equations to compute the hazard of infection over time, and applying standard survival analysis techniques to estimate natural history parameters of infection. This method has subsequently been extended to consider non-exponential distributions of key quantities such as the infectious period [13] and the impact of repeat testing in discrete time [16]. Variants of DSA have been used in multiple contexts, including Influenza A(H1N1) on a college campus [12], foot and mouth disease in the UK [13], Ebola in DRC [14, 17], and COVID-19 at a national level [13], at a state level [18, 19], in an individual prison [20], and on a college campus [16].

For simple models, such as the standard SIR, DSA involves solving a system of ordinary differential equations (ODEs) to compute the hazard of infection, which can be obtained using standard numerical solvers [12]. The assumption of exponentially distributed infectious periods in these models is, however, often unrealistic. Incorporating non-exponential distributions into DSA requires reformulating the transmission model using partial differential equations (PDEs) [13], which in turn typically require custom discretisation schemes. Together, these requirements may limit the adoption of DSA in practice.

Most studies using DSA to date have employed a continuous time, differential equation based model to compute the infection hazard. However, data such as symptom onset times and recovery times are normally recorded in the form of dates, thus continuous-time likelihoods must either interpolate the model’s solution or treat observed dates as approximations to continuous event times. However, data such as symptom onset and recovery times are typically recorded as dates. Thus, continuous-time likelihoods must either interpolate the model’s solution or treat observed dates as approximations of continuous event times. Wascher et al. [16] avoided this in their interval DSA, which uses a discrete-time Reed–Frost-type SEIR model to estimate prevalence and time-varying transmission from repeated SARS-CoV-2 testing in a closed population. Their model assumes a known population size and fixed sojourn-time distributions, and is fitted with a bespoke Metropolis–Hastings-within-Gibbs sampler that imputes individual infection and sojourn times. More recently, Roy et al. [15] addressed this challenge for temporally aggregated count data by deriving a closed-form marginal likelihood that treats daily or weekly incidence as interval-censored observations in continuous time. Here, we propose Discrete Dynamic Survival Analysis (DDSA), a discrete-time DSA for case line lists in which the population at risk can be either known or estimated, and extend it to non-geometric infectious period distributions and multiple groups, such as different age groups. The primary aim of this study is to make DSA easier to apply to line lists recorded by date. In discrete time, the likelihood is evaluated on the calendar days of the data, and such infectious periods are captured without potentially complex, model-specific numerical schemes, so that every model can be fitted with standard tools for Bayesian inference. We implement the framework in Julia using *Turing*.*jl* [21], and demonstrate inference on simulated and real-world line list data.

In DDSA, a recurrence map replaces the ODE and advances the epidemic state one step at a time, on the same time grid as the dated records. No solver is needed, and the map is compatible with forward-mode automatic differentiation, allowing the application of Markov chain Monte Carlo (MCMC) algorithms such as the No-U-Turn Sampler (NUTS) [22] directly. In discrete time, infectious periods that are not exponential can be modelled using discrete phase-type (DPH) distributions, which are specified by finite sub-stochastic matrices [23]. They include the negative binomial and distributions with finite support, and can approximate others. Their probability mass functions reduce to repeated matrix-vector products and are likewise differentiable with respect to the DPH parameters. Time-varying parameters, for example a contact rate that changes after an intervention, can enter the map as one value per step (*β*_*k*_), without changing how the likelihood is evaluated. The result is a complete inference pipeline for line list data, including flexible dwell-time distributions and multi-group population structure, that can be expressed concisely in a standard probabilistic programming language without a custom numerical framework.

To meet this aim, we (i) derive the discrete-time DSA log-likelihood and show that it converges to the continuous-time formulation as Δ*t* → 0; (ii) extend it to non-geometric infectious periods through two representations, a boxcar (infection-age) model and a discrete analogue of the ‘device of stages’; (iii) extend it to populations with multiple epidemiological groups; (iv) implement the full framework in Julia by fitting every model variant using the NUTS sampler in *Turing*.*jl*; and (v) assess what each part of the likelihood identifies, and how DDSA compares with continuous-time DSA, on synthetic data and on simulated outbreaks with known parameters. We demonstrate DDSA through applications to line list data from three real outbreaks: influenza A(H1N1) at a university campus [12], Ebola in the Democratic Republic of the Congo [14], and measles in Hagelloch, Germany, 1861 [24].

## 2 Methods

### 2(a) Dynamic survival analysis (DSA)

The Dynamic Survival Analysis (DSA) framework, as defined by [12], provides a formal link between mechanistic compartmental models and the statistical theory of survival analysis. By interpreting the transition flows of a mean-field epidemic model as individual-level hazards, DSA enables the construction of product-form likelihood functions from line-list data. Throughout, we write such likelihoods multiplicatively as ℒ and the corresponding log-likelihood additively as *ℓ* = log ℒ; the large population assumption results in the likelihoods for each individual in the line list being independent, while keeping sampling variation from a limited number of individuals in a line list.

In a closed, well-mixed population with *n* initially susceptible individuals (where *n* is assumed to be large), the standard susceptible-infected-removed (SIR) dynamics are governed by:

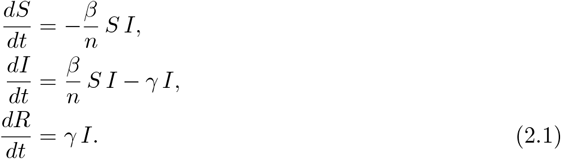

DSA recasts this ODE model as a set of survival-analytic objects describing the infection and recovery times of a single individual. Integrating Eq. (2.1) yields the equivalent integral representation for the proportions *s*(*t*) = *S*(*t*)*/n* and *i*(*t*) = *I*(*t*)*/n*:

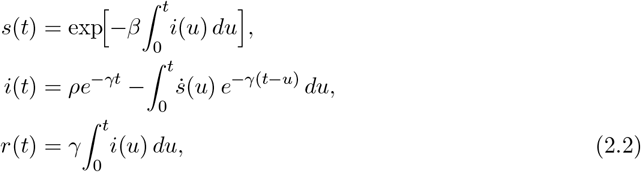

where *ρ* = *i*(0) is the number of initial infectives as a fraction of *n*. Scaling by *n* makes *s*(0) = 1, so *s*(*t*) is the probability that an initially susceptible individual has not been infected by time t, the survival function on which DSA builds its likelihood. Here, the cumulative hazard is 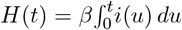, and the instantaneous hazard (force of infection) is *h*(*t*) = *βi*(*t*). Within this framework, the mean-field ODE acts as a Chapman–Kolmogorov equation for a 3-state Markov chain; thus, DSA provides a mathematically consistent mapping from a deterministic compartmental model to an individual-level stochastic process. This correspondence becomes exact as the population grows large. As *N* → ∞ , the scaled prevalence *I*(*t*)*/N* converges to the deterministic solution *i*(*t*) by the law of large numbers, so at finite *N* the individual-level hazards in Eq. (2.2) are an approximation whose error shrinks as *N* increases. In simulation, KhudaBukhsh et al. [12] found this approximation accurate for populations as small as *N* ∼ 10^2^, with the initial infectious fraction *ρ* the most sensitive parameter at small *N* .

#### 2(a)(i) Exponential infectious period

Treating (*S, I, R*) as survival objects yields a product-form likelihood for any random sample of infection and/or recovery times, accommodating censoring. For a sample of *K* newly infected individuals with infection times *t*_*i*_ and recovery intervals *w*_*i*_, and *M* initially infectious individuals with recovery intervals *ϵ*_*j*_, the log-likelihood is [12]:

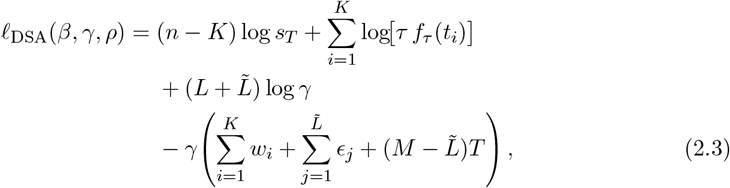

where *T* is the end of the observation window, and *s*_*T*_ = *s*(*T*) is the susceptible fraction at that time. The normalised infection-time density is *f*_*τ*_ (*t*) = *βτ*^−1^*i*_*t*_*s*_*t*_, and *τ* = 1 − *s*_*T*_ is the attack rate. The number of newly infected individuals who recover within [0, *T*] is *L*, and the number of the *M* initially infectious individuals who recover within [0, *T*] is 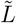. Right-censored recovery times enter the final term as *w*_*i*_ = *T* − *t*_*i*_ or *ϵ*_*j*_ = *T* for the remaining *K* − *L* and 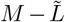 individuals, respectively.

#### 2(a)(ii) Non-exponential infectious periods

The assumption of an exponentially distributed infectious period is often biologically unrealistic. Di Lauro et al. [13] extended Eq. (2.3) to accommodate any absolutely continuous distribution for the infectious period *W* , governed by parameters *ϕ*. In Eq. (2.3), the last two terms collect the recovery contributions. Each of the 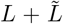 observed recovery intervals *w* contributes log(*γe*^−*γw*^), and each censored interval Δ (*T* − *t*_*i*_, or *T* for an initial infective) contributes log *e*^−*γ*Δ^. Di Lauro et al. replaced the exponential density *γe*^−*γw*^ by an arbitrary density *f*_*W*_ (· | *ϕ*), and a censored interval then contributes the survivor *S*_*W*_ (Δ | *ϕ*) in place of *e*^−*γ*Δ^.

We introduce notation used throughout: *ℐ*_R_ and *ℐ*_NR_ denote the sets of newly infected individuals who recover within [0, *T*] and those still infectious at *T* , respectively; 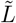 is the number of initially infectious individuals (*M* in total) who recover; *w*_*i*_ and *ϵ*_*j*_ are the respective recovery intervals; and *s*_*T*_ = *S*(*T*)*/n* is the susceptible fraction at the end of observation. The full likelihood factorises as [13]:

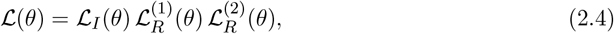

where ℒ_*I*_ uses infection times via the normalised density *f*_*τ*_ (*t*); 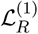 uses observed infectious-period durations *w*_*i*_ with *f*_*W*_ (· | *ϕ*); and 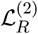 handles cases where only recovery times are recorded, requiring a convolution of the infection-time density *f*_*T,θ*_ (conditional on infection by *T*) and the infectious-period density 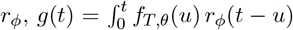 *du* with no closed form. As with the other terms, these records can be a random sample of the individuals whose infection times are unknown.

Implementing Eq. (2.4) generally requires tracking the infection-age distribution *y*_*I*_(*t, s*) via an age-structured PDE and a custom numerical discretisation scheme [13]. This underlying PDE complexity motivates the discrete-time reformulation described in the next section.

### 2(b) Discrete dynasmic survival analysis (DDSA)

The transition from a continuous-time ODE to a discrete-time function map can facilitate an efficient inference pipeline. By defining the epidemic state on a grid *t*_*k*_ = *k*Δ*t*, we obtain the following update equations:

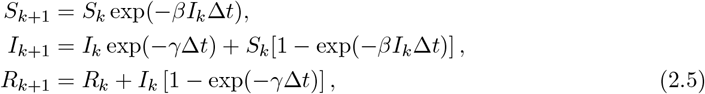

This formulation avoids the need for an ODE solver within the likelihood evaluation loop. The map matches the discrete-time SIR model of Brauer et al. [25], and the multiplicative escape probability, exp(− *βI*_*k*_Δ*t*), keeps *S*_*k*_ non-negative, consistent with each host being infected at most once [26]. In the limit Δ*t* → 0, the map converges to the continuous-time SIR model. As each update multiplies the states by probabilities, *S*_*k*_, *I*_*k*_ and *R*_*k*_, remain non-negative for any Δ*t*. The susceptible trajectory *S*_*k*_ maintains its interpretation as a survival function, so the survival-analytic reinterpretation established by [12] carries over to the discrete case (Table 1).

DDSA replaces each element of the continuous-time likelihood (2.4) with a discrete analogue: continuous distributions *f*_*W*_ , *S*_*W*_ are replaced by discrete phase-type (DPH) families (Section 2(b)(i)), and the likelihood retains the three-component structure *ℓ*_*I*_ + *ℓ*_*R*_ + *ℓ*_constr_. Because every step reduces to elementary differentiable operations on the parameters, the log-posterior is differentiable via forward-mode automatic differentiation, and inference methods can be applied directly.

**Table 1:**
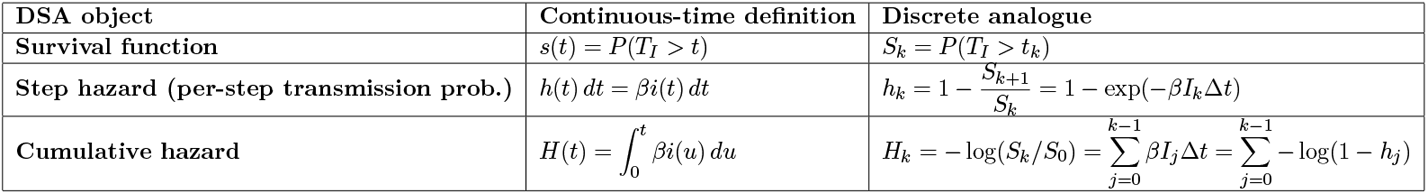
Comparison of continuous-time definitions and discrete analogues in DSA.

| **DSA object** | **Continuous-time definition** | **Discrete analogue** |
|---|---|---|
| **Survival function** | *s*(*t*) = *P*(*T*<sub>*I*</sub> > *t*) | *S*<sub>*k*</sub> = *P*(*T*<sub>*I*</sub> > *t*<sub>*k*</sub>) |
| **Step hazard (per-step transmission prob.)** | *h*(*t*) *dt* = *βi*(*t*) *dt* | $h_k = 1 - \frac{S_{k+1}}{S_k} = 1 - \exp(-\beta I_k \Delta t)$ |
| **Cumulative hazard** | $H(t) = \int_0^t \beta i(u)\, du$ | $H_k = -\log(S_k/S_0) = \sum_{j=0}^{k-1} \beta I_j \Delta t = \sum_{j=0}^{k-1} -\log(1-h_j)$ |

The infection time *T*_*I*_ is defined as a discrete random variable with probability mass function

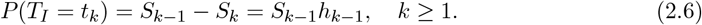

where *T*_*I*_ is the infection time of a randomly chosen member of the whole population, with *T*_*I*_ = *t*_0_ for the initial infectives, and *S*_*k*_ = *P* (*T*_*I*_ *> t*_*k*_) is the discrete survival function. The step hazard

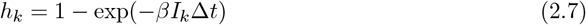

is the per-step transmission probability induced by an underlying Poisson process: if a susceptible individual experiences infectious contact at rate *βI*_*k*_ over the interval [*t*_*k*_, *t*_*k*_ + Δ*t*), then the probability of at least one transmission event in that interval is 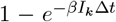. The corresponding cumulative hazard is

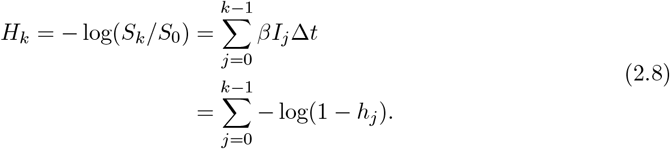

For small Δ*t, h*_*k*_ = *βI*_*k*_Δ*t* + *O* (Δ*t*^2^), recovering the continuous-time hazard *βI*(*t*) *dt* in the limit Δ*t* → 0. This discrete-time formulation accommodates any reporting cadence (e.g., daily or weekly observations) by choosing Δ*t*. Consequently, no smoothing or interpolation of the epidemic curve is required. DDSA keeps the individual-level structure of the continuous-time likelihood, in which each case contributes its own infection time and recovery interval, while matching data recorded by day.

#### 2(b)(i) Inference

Taking logs of the factorised continuous-time likelihood in Eq. (2.4), the DDSA log-likelihood is partitioned into three additive components:

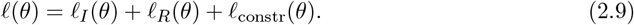

##### Infection component

The discrete-time SIR map generates a PMF **f** = (*f*_1_, … , *f*_*T*_) across the infection steps, where each element is defined as *f*_*k*_ = (*S*_*k*−1_ − *S*_*k*_)*/τ* and the attack rate is *τ* = *S*_0_ − *S*_*T*_ = 1 − *ρ* − *S*_*T*_ , the new infections as a fraction of the whole population, with initial conditions *S*_0_ = 1 − *ρ* and *I*_0_ = *ρ*. For a sample of *K* observed infection times summarised as daily counts **y** = (*y*_1_, … , *y*_*T*_), where *y*_*k*_ is the number of cases infected in step *k*, the likelihood contribution is:

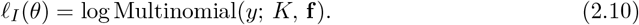

##### Recovery component

For a geometrically distributed infectious period with per-step recovery probability *p*_*γ*_ = 1 − *e*^−*γ*Δ*t*^, the probability mass function *P* (*W* = *n*) and survivor function *P* (*W > n*) are defined as:

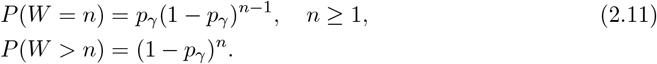

Using the observation-set notation established in Section 2(a)(ii), each observed recovery interval **δ**_*i*_ (the days from infection to recovery of case *i*) enters the likelihood through the geometric PMF, while right-censored intervals, which represent individuals still infectious at the end of the observation window, enter through the survivor function:

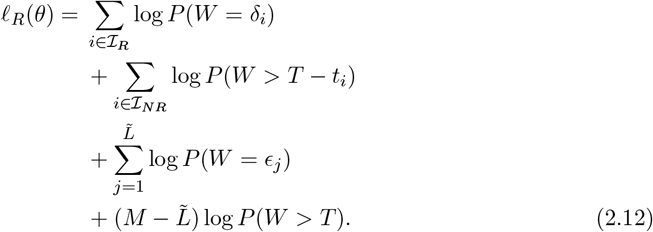

##### Final-size constraint

The total infection count *K* in the whole population of *N* individuals contributes a binomial term to the likelihood, representing the probability of the observed epidemic scale given the model-derived attack rate:

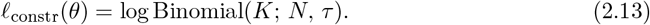

Here *N* includes the initial infectives, and the final-size term treats each individual’s probability of not being newly infected as 1 − *τ* = *ρ* + *S*_*T*_ .

In the analyses presented here, inference is based on the compact form in Eqs. (2.10)–(2.13). In the synthetic data experiments in Section 2(c)(i), all recovery intervals are fully observed, so the recovery contribution reduces to independent PMF terms over the *M* + *K* observed durations. In the H1N1 analysis, right-censored recoveries contribute survivor terms, while in the Ebola analyses an additional convolution term is introduced for hospitalisation-only records, marginalising over unknown onset times while retaining the same DDSA infection and final-size structure. Cases with a missing recovery contribute only to *ℓ*_*I*_ and *ℓ*_constr_, which assumes that recoveries are missing at random. The compact three-component decomposition therefore serves as the core DDSA likelihood, with dataset-specific missing-data terms added when required by the observation process.

As Δ*t* → 0 with *p*_*γ*_ = 1 − *e*^−*γ*Δ*t*^ ≈ *γ*Δ*t*, the compact DDSA likelihood components converge to their continuous-time analogues. Specifically, the geometric survival function (1 − *p*_*γ*_)^*n*^ converges to the exponential survivor *e*^−*γn*Δ*t*^ and the discrete PMF *p*_*γ*_(1 − *p*_*γ*_)^*n*−1^ converges to the exponential density *γe*^−*γn*Δ*t*^Δ*t*, recovering the exponential-recovery likelihood of KhudaBukhsh et al. [12]. This convergence implies that the DDSA framework recovers the exponential recovery likelihood of KhudaBukhsh et al. [12] as the time step approaches zero.

For non-geometric infectious periods, the geometric PMF and survivor function in Eq. (2.11)–(2.12) are replaced by those of the appropriate discrete phase-type (DPH) distribution as defined in Eq. 2.26. This replacement is modular, requiring no structural changes to the infection or final-size components of the likelihood. More broadly, the same three-component likelihood applies to any difference-equation compartmental model that yields a susceptible trajectory 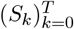 : extensions such as SEIR, time-varying transmission, or additional compartments enter only through the recurrence map and require no change to the infection-time, recovery, or final-size terms. The complete inference procedure, which integrates these components into a unified Bayesian framework, is summarised in Table 2.

**Table 2:**
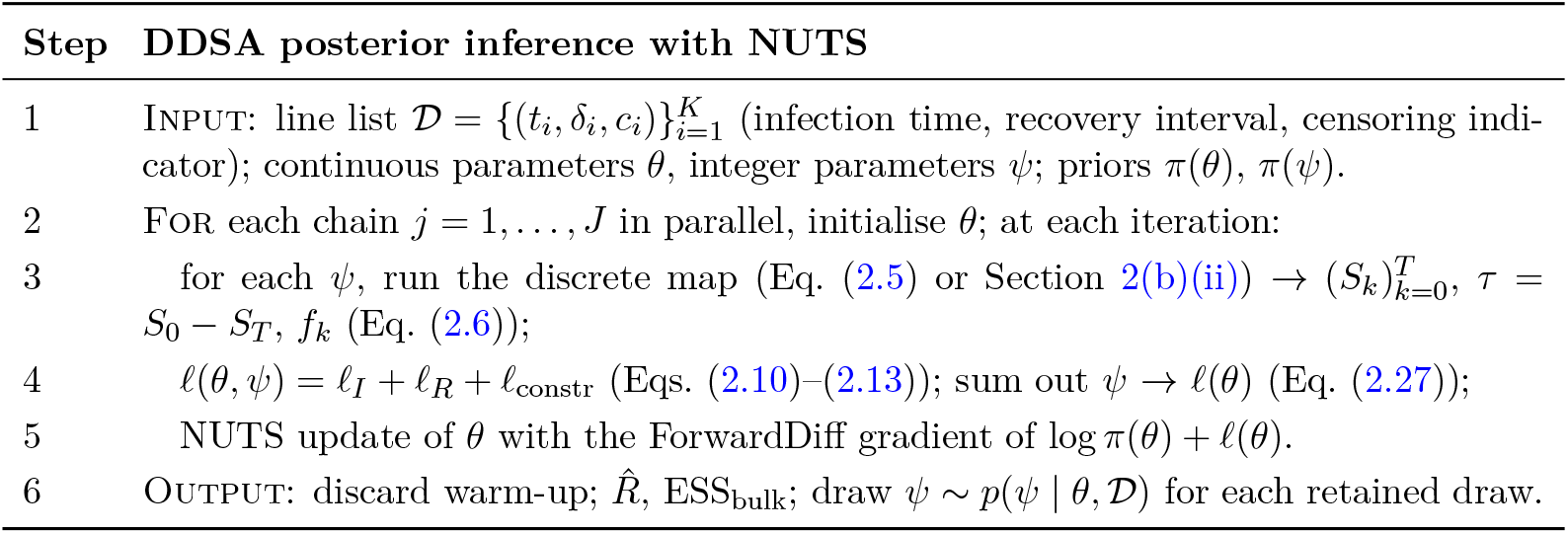
DDSA inference algorithm. The discrete map (step 3) replaces the ODE solver required by continuous DSA. The log-posterior is differentiable in *θ*, and integer parameters are summed out, so every model is sampled with NUTS alone. Settings in Section 2(d).

| **Step** | **DDSA posterior inference with NUTS** |
|---|---|
| 1 | Input: line list $\mathcal{D} = \{(t_i, \delta_i, c_i)\}_{i=1}^{K}$ (infection time, recovery interval, censoring indi-cator); continuous parameters θ, integer parameters ψ; priors π(θ), π(ψ). |
| 2 | For each chain *j* = 1, . . . , *J* in parallel, initialise θ; at each iteration: |
| 3 | for each ψ, run the discrete map (Eq. (2.5) or Section 2(b)(ii)) → $(S_k)_{k=0}^{T}$, τ = *S*<sub>0</sub> − *S*<sub>*T*</sub>, *f*<sub>*k*</sub> (Eq. (2.6)); |
| 4 | ℓ(θ, ψ) = ℓ<sub>*I*</sub> + ℓ<sub>*R*</sub> + ℓ<sub>constr</sub> (Eqs. (2.10)–(2.13)); sum out ψ → ℓ(θ) (Eq. (2.27)); |
| 5 | NUTS update of θ with the ForwardDiff gradient of log π(θ) + ℓ(θ). |
| 6 | Output: discard warm-up; $\hat{R}$, ESS<sub>bulk</sub>; draw ψ ∼ $p(\psi \mid \theta, \mathcal{D})$ for each retained draw. |

#### 2(b)(ii) Model formulations

The base SIR map assumes a geometric infectious period, the discrete analogue of an exponential one, so recovery is memoryless: the chance of recovering on a given day does not depend on how long an individual has been infected. For many pathogens this is unrealistic. Their infectious periods are less variable, with probability concentrated away from zero and a lower coefficient of variation than the geometric [27]. Modelling this lower variability produces more realistic epidemic trajectories. Other discrete-time epidemic models have also used non-geometric stage durations, for example, to study control measures [28] or generation times [29]. We accommodate such non-geometric infectious periods through two complementary discrete-time representations. The Boxcar model tracks infection age and admits any discrete distribution through an age-specific hazard sequence. The Generalized Linear Chain Trick (GLCT) instead chains staged compartments to approximate any distribution in the discrete phase-type family, needing more stages the less variable the distribution is. Both change how the infected leave the infectious class in the map, and therefore *τ* and **f** , which modify the recovery term *ℓ*_*R*_ of the likelihood. Meanwhile, the form of the infection-time (*ℓ*_*I*_) and final-size (*ℓ*_constr_) components remain unchanged.

The two models use different DPHs. The Boxcar model has one stage for each day of infection age, up to *K*_age_, where it is truncated. This allows it to follow any hazard sequence, whether estimated from data or derived from a parametric distribution. The GLCT map can represent any DPH, with one compartment per stage, and needs no truncation. For the same infectious-period distribution, the two models produce the same susceptible trajectory, except for truncation at *K*_age_, which is negligible when *P* (*W > K*_age_) is small.

The DDSA framework supports three complementary mechanistic model formulations, each corresponding to a different representation of the infectious period distribution:

##### Discrete SIR map

The base formulation iterates the SIR map in Eq. (2.5) over the observation window to generate the susceptible trajectory 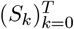. The normalised infection-time PMF is defined as *f*_*k*_ = (*S*_*k*−1_ − *S*_*k*_)*/τ* with attack rate *τ* = *S*_0_ − *S*_*T*_ . In this configuration, the infectious period follows a geometric distribution with a per-step recovery probability *p*_*γ*_ = 1 − *e*^−*γ*Δ*t*^. The Boxcar and GLCT formulations, described below, extend this framework to accommodate non-geometric infectious periods without changing the form of the infection and final-size components of the likelihood.

##### Boxcar model

Let *S*_*k*_ denote the susceptible fraction at day *k, I*^(*a*)^ denote the infectious fraction in infection-age class *a* (*a* = 1, … , *K*_*age*_) , and *R*_*k*_ denote the removed fraction at day *k*. Here, *K*_*age*_ is the maximum infection age explicitly tracked. New infections enter class *a* = 1, then advance one class per time step if they do not recover. In practice, *K*_*age*_ is chosen sufficiently large that the survival probability beyond *K*_*age*_ is negligible.

Any discrete recovery-time distribution is characterised through its survival function *G*(*a*) = *P* (*W > a*), for *a* ∈ {0, 1, … }, and the corresponding age-specific hazard:

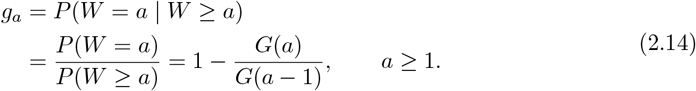

The Boxcar formulation allows arbitrary discrete infectious-period distributions by explicitly tracking infection age, providing a flexible alternative to the memoryless standard SIR map [28]. The complete system of update equations governing the Boxcar model is given by:

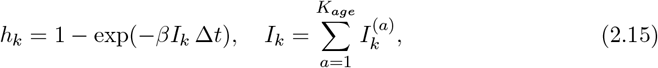

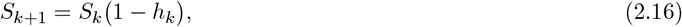

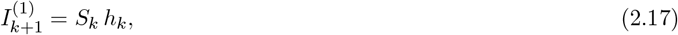

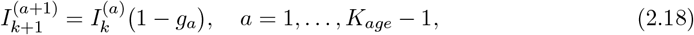

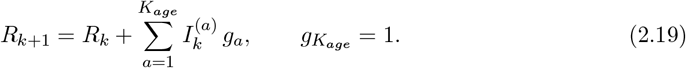

The scheme is conservative, satisfying 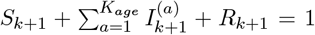 at every step. The per-step transmission probability *h*_*k*_ = 1 − exp(− *βI*_*k*_Δ*t*) is the exact discrete-time hazard, consistent with the base SIR map in Eq. (2.5).

When the recovery-time distribution is geometric and *K*_*age*_ is chosen large enough that terminal truncation is negligible over the observation window, the ageing equations simplify to 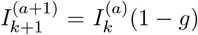 for *a* = 1, … , *K*_*age*_ − 1 with *g*_*a*_ ≡ *g*. Summing over the age index *a* then shows that the aggregate infectious class evolves according to *I*_*k*+1_ = *I*_*k*_(1 − *g*)+*S*_*k*_ *h*_*k*_, recovering the single-compartment DDSA formulation away from the terminal boundary. Consequently, the Boxcar representation reduces to the original difference equations whenever the recovery distribution is geometric.

Setting 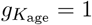 limits the infectious period to *K*_age_ steps, so *K*_age_ must be large enough for the truncated tail to be negligible. As Δ*t* → 0, the per-step hazard satisfies *g*_*a*_ ≈ *h*_*W*_ (*a*Δ*t*) Δ*t*, where *h*_*W*_ is the hazard of the continuous-time infectious period. The Boxcar map is then a discretisation, with equal steps in time and infection age, of the age-structured PDE that Di Lauro et al. use for non-exponential infectious periods (Section 2(a)(ii)):

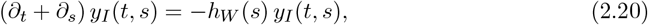

where *y*_*I*_(*t, s*) is the density of infectives with infection age *s* at time *t*.

##### Discrete-time generalized linear chain trick (GLCT map)

The Linear Chain Trick (LCT) replaces the single infectious compartment *I* with *k* exponential sub-stages, each governed by the same rate *γ*, resulting in an Erlang-distributed infectious period within an ODE system. Hurtado and Kirosingh [30] extended this to the Generalized Linear Chain Trick (GLCT), which allows for varying rates per stage, thereby enabling the representation of any distribution in the phase-type family (e.g., exponential, Erlang, Coxian, or hyperexponential). The theoretical foundation is the “device of stages” by Cox and Miller [31], which builds non-exponential waiting times from memoryless stages in series or parallel. A DPH with *r* stages and mean *m > r* steps has CV^2^ ≥ 1*/r* − 1*/m*, attained by the negative binomial [23, 32]. While these elementary stages are exponential in continuous time, they are geometric in the discrete-time formulation. The update rule for a single stage s is:

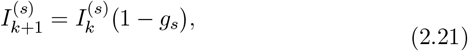

where *g*_*s*_ ∈ (0, 1] represents the per-step progression probability from stage *s*. In continuous time, the GLCT dynamics are governed by 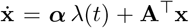, where **A** is the sub-generator matrix and ***α*** is the initial phase distribution. The DPH analogue used in this study is obtained stage by stage. At each stage, an exit rate *λ*_*s*_ is retained for the next step with probability 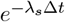 or passed on with probability 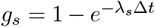. This passed probability is then allocated to different destinations based on the rates in the matrix **A**. These probabilities form a substochastic matrix **T**, such that Eq. (2.21) takes the matrix form **x**_*k*+1_ = **T**^T^**x**_*k*_ + ***α*** *h*_*k*_*S*_*k*_. This construction allows at most one stage transition per step, ensuring that each stage remains geometric. Another DPH comes from sampling the continuous-time chain every Δ*t*, represented by **T** = *e*^**A**Δ*t*^. This approach allows multiple transitions within a single step [33]. All analyses in this study used serial DPHs constructed stage by stage.

For *r* identical serial stages each with progression probability *g*, the infectious period *W* (in steps of length Δ*t*) is the sum of *r* independent geometric(*g*) variables, yielding a negative-binomial distribution:

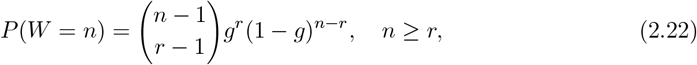

with mean *E*[*W*] = *r/g* steps. As Δ*t* → 0 with *g* = 1 − *e*^−*λ*Δ*t*^, *W* Δ*t* converges to the Erlang distribution with *r* stages and rate *λ* (Section 2(b)(i)).

With force-of-infection *h*_*k*_ defined as above, the governing difference equations for the staged progression model are:

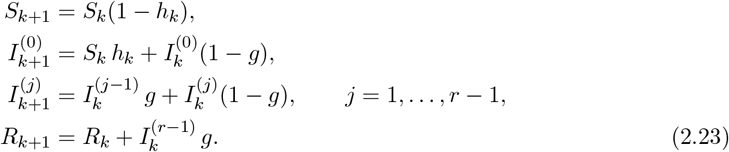

When *r* = 1, these equations reduce to the base map in Eq. (2.5). For *r >* 1, they are the equations of the discrete staged-progression model [25], the case applied in the analyses presented here.

To capture heterogeneity in the infectious period, such as the distinction between mild and severe clinical pathways, individuals may branch into different trajectories upon infection. Let *q*_*b*_ be the probability of entering branch *b*, where **∑***q*_*b*_ = 1. Each branch *b* contains *r*_*b*_ serial stages with progression probabilities 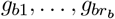 . The aggregate survival function is expressed as a convex mixture:

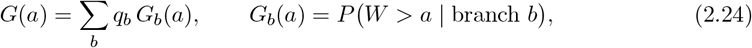

where each *G*_*b*_ follows the negative binomial form derived previously when the progression probabilities within branch *b* are equal, and is otherwise the survivor function of a sum of geometric variables with different probabilities. By selecting an appropriate number of branches and stages, this formulation can represent acyclic DPHs [23], including the geometric, negative-binomial and Coxian families. The corresponding governing difference equations for this branched system are:

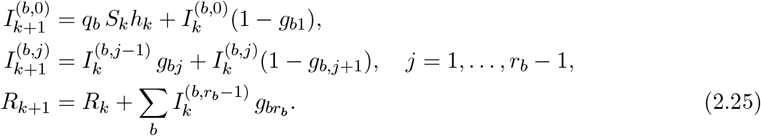

The total number of infectives at day *k* is 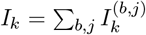 , while the susceptible population evolves exactly as in the previous formulations. Although the analyses in this paper used only serial DPHs, the GLCT map in the accompanying code also accepts non-serial DPHs.

The device-of-stages formulation may require fewer compartments than the boxcar model and avoids the need to truncate the infection-age distribution. However, it requires committing to a specific parametric family: the number of stages *r* and their associated progression probabilities must be specified or inferred, rather than being determined directly from the data. In contrast, the Boxcar representation accommodates any discrete distribution (empirical or parametric) simply by supplying the hazard sequence *g*_*a*_. When a parametric family such as the negative binomial or Erlang is adopted, the two representations are equivalent and yield identical state trajectories for the same parameter values. In such cases, the device-of-stages is preferred for its compact state vector and its compatibility with gradient-based samplers like NUTS, which benefit from the smooth, differentiable parameterisation of the phase-type transition matrix.

### 2(c) Datasets

#### 2(c)(i) Synthetic data

Synthetic line lists were generated from the discrete GLCT map with four geometric stages (*r* = 4) at Δ*t* = 1 d, each discretising an exponential stage with rate *λ* = 1 d^−1^ (continuous-time mean 4 d), with *β* = 0.5 day^−1^ and *N* = 99. The values to recover were *R*_0_ = 3.16 and a mean infectious period of 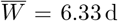. This mean exceeds the continuous-time mean because a geometric stage lasts longer on average than the exponential stage it discretises, and the two agree as Δ*t* → 0 (Supplementary Section S1.4). The difference matters when comparing discrete-time and continuous-time quantities. For example, fixing a continuous-time removal rate at 1*/*4 d^−1^ biases *R*_0_ (Section 3(b)).

#### 2(c)(ii) Historical case studies

We used three historical outbreak line lists, each chosen to illustrate a different aspect of the DDSA framework (Table 3). All analyses assumed that the recorded cases were a random sample of infections in the population at risk, so whether a case was recorded did not depend on its infection date or infectious period [12, 13]. Unrecorded cases then lower the estimated population size *N* , which is why *N* is read as an effective size, but leave the infection-time and recovery terms unchanged.

**Table 3:**
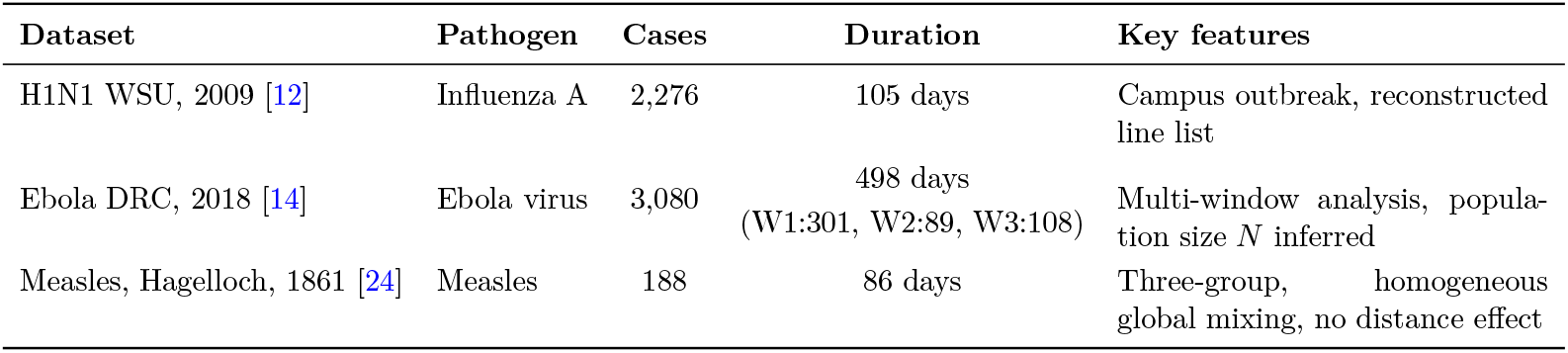
Historical outbreak datasets used to evaluate DDSA.

| **Dataset** | **Pathogen** | **Cases** | **Duration** | **Key features** |
|---|---|---|---|---|
| H1N1 WSU, 2009 [12] | Influenza A | 2,276 | 105 days | Campus outbreak, reconstructed line list |
| Ebola DRC, 2018 [14] | Ebola virus | 3,080 | 498 days<br>(W1:301, W2:89, W3:108) | Multi-window analysis, population size *N* inferred |
| Measles, Hagelloch, 1861 [24] | Measles | 188 | 86 days | Three-group, homogeneous global mixing, no distance effect |

##### H1N1 WSU, 2009

The source dataset contains 2,276 rows. KhudaBukhsh et al. reconstructed individual infection times from daily counts. The infection times are spread within each day, with 11 initially infected cases at time 0, matching the initial number infected in an earlier model of this outbreak [34]. They generated recovery times by sampling from an exponential distribution with mean *γ*^−1^ = 5.5 days, a value fixed from infectious-period estimates rather than observed [12]. Of the sampled recovery times, 13 fell after the end of the observation window (day 104) and were treated as right-censored.

##### Ebola DRC, 2018

The 2018 Ebola outbreak comprised three successive waves, each analysed independently, together covering the period from the first recorded onset to 12 September 2019 (Wave 1 to 27 February 2019, Wave 2 to 27 May 2019, and Wave 3 to 12 September 2019). The outbreak database described by Vossler et al. [14] held 3,117 case records (confirmed and probable) recorded between 3 May 2018 and 12 September 2019. Of these, 37 records with neither an onset nor a hospitalisation date were excluded, leaving 3,080 cases (Waves 1–3: 907, 1,104 and 1,069). Some of the remaining records were missing either the onset or the hospitalisation date. The hospitalisation date was used as a proxy for removal from transmission. Section 3(d) describes how each record type enters the likelihood.

##### Measles, Hagelloch, 1861

The line list identifies three epidemiological classes (pre-school, younger school, and older school students) with individual prodrome-onset and rash-onset dates for all 188 cases [35]. Infectious-period durations **δ**_*i*_ = (rash date − prodrome date) + 4 days follow Neal and Roberts [36] with *d*_0_ = 3 and *d*_1_ = 1. Each child is taken to be infectious from *d*_1_ day before the onset of prodromal symptoms until *d*_0_ days after the rash appears. The mean infectious period of approximately 8 days and coefficient of variation of 0.22 indicate that a NegBin DPH is more appropriate than a geometric model.

### 2(d) Implementation

All analyses were implemented in the Julia programming language [37] using the probabilistic programming framework *Turing*.*jl* [21] for Bayesian inference. The code and notebooks for each analysis are archived on Zenodo [38]. DDSA is also available as a Julia package DiscreteDynamicSurvivalAnalysis.jl [39], whose vignettes reproduce these analyses.

The analyses were implemented by evaluating the discrete recurrences from Section 2(b) directly within the likelihood, with continuous transition rates converted to per-step event probabilities via expressions of the form *p* = 1 − *e*^−*λ* Δ*t*^. As a result, the log-posterior remains differentiable in the continuous parameters, allowing the use of gradient-based inference in *Turing*.*jl*.

Priors and bounds for every analysis are listed in Supplementary Section S1. Integer-valued parameters had discrete uniform priors and were summed out of the likelihood.

Uniform priors were adopted in settings where epidemiologically motivated bounds could be specified from the data context. For transmission and removal rates, upper bounds were chosen to exceed plausible values for the pathogen and population, while lower bounds preclude zero or near-zero rates that are inconsistent with an observed outbreak. For initial infected fractions, a Beta prior concentrated near zero reflects the expectation of a small founding seed. These uniform priors assign equal mass within their support and do not penalise values near the bounds, which can affect posterior shape when data are sparse. Sensitivity to the prior bounds is examined in Supplementary Section S2.3. However, log-normal or half-normal alternatives for rate parameters are a direction for future work.

All DDSA analyses used the same sampler settings: NUTS with four chains of 2,000 post-warm-up draws after 1,000 warm-up iterations, with 4,000 draws for the NegBin DPH and Boxcar fits of Scenario B. Chains were started from jittered data-based values, as chains started from prior draws occasionally became trapped in low-density regions in replicated simulations.

The convergence targets were 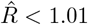 and ESS_bulk_ ≥ 100 per parameter [40]. The count-based comparators of Section 3(b) were sampled with longer warm-up and started from the prior. Full sampler settings and starting values are given in Supplementary Section S5.1. Gradients were computed with ForwardDiff [41] via the default *Turing*.*jl* automatic differentiation backend.

To handle non-exponential infectious periods, we implemented a self-contained phase-type module in the accompanying code, which integrates continuous phase-type (CPH) and discrete phase-type (DPH) families within the standard *Distributions*.*jl* interface [42]. The distinction between the two parametrisations is computationally important. A CPH with generator matrix **A** has density *f*_*W*_ (*t*) = ***α***^T^*e*^**A***t*^ **q**, where **q** = − **A1**. Evaluating the matrix exponential at each observed recovery time is computationally expensive and often challenging for scalar automatic differentiation. In contrast, a DPH with a sub-stochastic transition matrix **T** has a probability mass function defined as:

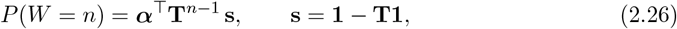

This requires only matrix products and no matrix exponential. For the serial DPHs used in this paper, the module identifies the stage structure and evaluates Eq. (2.26) in closed form using Eq. (2.22). Other DPHs are evaluated using the matrix form. A DPH is constructed stage by stage from a CPH generator, as described in Section 2(b)(ii).

Integer-valued parameters, namely the DPH phase count *r* and the population sizes *N* (or *N*_0_, *N*_1_, *N*_2_ in the multi-group model), are discrete, whereas NUTS requires a differentiable target. We therefore summed them out of the likelihood exactly over their finite support, under discrete uniform priors, so that every model was sampled with NUTS alone. With *ψ* denoting the integer parameters,

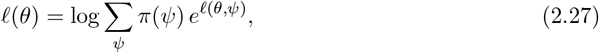

When a population size enters only the Binomial final-size term, the sum is a single pass over its support, with no further evaluations of the recurrence map. When a parameter changes the map or the recovery distribution (the phase count *r*, or *N*_2_ in the Hagelloch model, which also sets the seed *ρ*_2_ = 1*/N*_2_), each value requires one evaluation of the map, and parameters that vary together are summed over their joint grid. After sampling, the summed-out parameters were drawn, once per retained draw, from their exact conditional posterior, *p*(*ψ* | *θ, D*) ∝ *π*(*ψ*) *eℓ*(*θ,ψ*).

## 3 Results

### 3(a) Data availability: inference from partial line lists

The contribution of each likelihood term was quantified by fitting DDSA to the synthetic dataset of Section 2(c)(i) under three data scenarios: (A) the full line list (*ℓ*_*I*_ +*ℓ*_*R*_ +*ℓ*_constr_); (B) infection times only (*ℓ*_*I*_ + *ℓ*_constr_); and (C) recovery intervals only (*ℓ*_*R*_). In Scenarios A and B, GLCT and Boxcar models with a NegBin DPH and a geometric model were fitted. In Scenario C, the GLCT and Boxcar models are identical, so one NegBin DPH model and a geometric model were fitted. The phase count *r* was summed out of the likelihood (Supplementary Section S1.4).

Results are summarised in Table 4 and Figure 1. Recovery intervals alone (Scenario C) recovered the mean infectious period, with *r* at its cap of 4, which equalled the true value here, while *β* was not identified. A geometric fit to the same intervals produced a similar mean with a wider range, but its predicted interval distribution decreased consistently. It missed the observed peak at 6–7 d and placed about 37 of the 92 intervals at 1–3 d, where none were observed (Figure 2, bottom row). With infection times only (Scenario B), *R*_0_ was constrained, with intervals that included the true values for the GLCT and Boxcar fits. However, their upper limits reflected the U(0.5, 5) prior (Supplementary Section S2). The infectious-period shape remained uncertain: the posterior of *r* was spread over *r* = 2–7 and the mean-infectious-period interval was wide. The full line list (Scenario A) returned narrow posteriors for all parameters, with the GLCT and Boxcar posteriors overlapping closely.

**Table 4:**
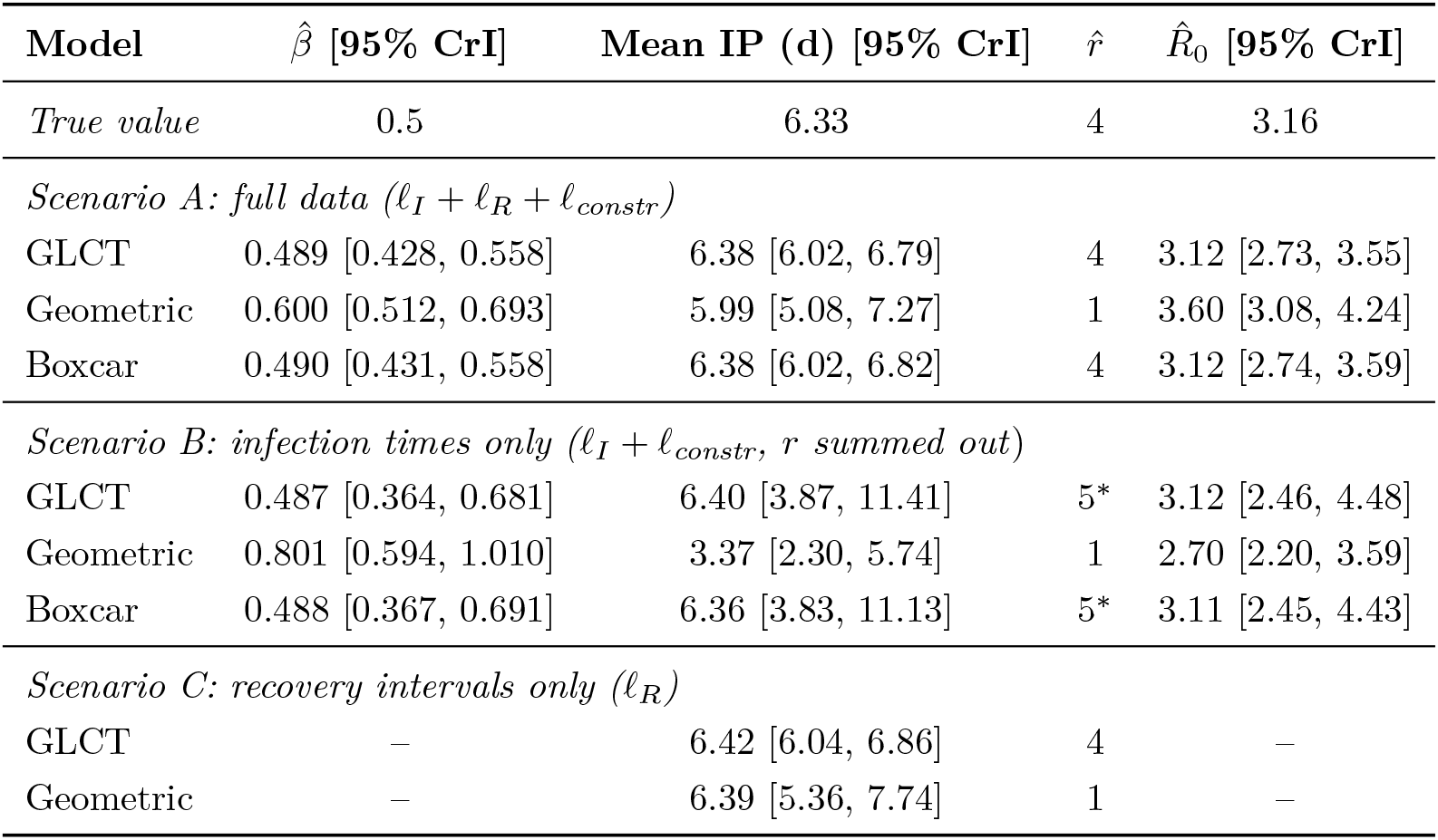
Data-availability scenarios for the synthetic study. Posterior summaries for the NegBin discrete phase-type (DPH) infectious period, fitted via the GLCT and Boxcar maps (rows ‘GLCT’ and ‘Boxcar’), and for a misspecified geometric model (*r* ≡ 1), across three observation scenarios: A (full line list), B (infection times only, *r* summed out of the likelihood), and C (recovery intervals only). Bracketed ranges are 95% credible intervals (CrI). Dashes denote parameters not identified by the available data. All entries are posterior medians; *R*_0_ is computed draw by draw as 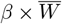 (and sampled directly by the GLCT and Boxcar models in Scenario B), then summarised. True values are those of the discrete map that generated the data (Section 2(c)(i)), with 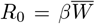. ^*^Posterior median of *r*, whose posterior is spread over *r* = 2–7 (GLCT: *P* (*r* = 4) = 0.20, *P* (*r* = 5) = 0.21). All fits use four chains of 2,000 post-warm-up draws (4,000 for the GLCT and Boxcar fits of Scenario B), started from jittered data-based values, with 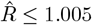 and bulk ESS ≥ 1,238 for every model.

| **Model** | **$\hat{\beta}$ [95% CrI]** | **Mean IP (d) [95% CrI]** | $\hat{r}$ | **$\hat{R}_0$ [95% CrI]** |
|---|---|---|---|---|
| *True value* | 0.5 | 6.33 | 4 | 3.16 |
| *Scenario A: full data (ℓ<sub>I</sub> + ℓ<sub>R</sub> + ℓ<sub>constr</sub>)* |  |  |  |  |
| GLCT | 0.489 [0.428, 0.558] | 6.38 [6.02, 6.79] | 4 | 3.12 [2.73, 3.55] |
| Geometric | 0.600 [0.512, 0.693] | 5.99 [5.08, 7.27] | 1 | 3.60 [3.08, 4.24] |
| Boxcar | 0.490 [0.431, 0.558] | 6.38 [6.02, 6.82] | 4 | 3.12 [2.74, 3.59] |
| *Scenario B: infection times only (ℓ<sub>I</sub> + ℓ<sub>constr</sub>, r summed out)* |  |  |  |  |
| GLCT | 0.487 [0.364, 0.681] | 6.40 [3.87, 11.41] | 5<sup>*</sup> | 3.12 [2.46, 4.48] |
| Geometric | 0.801 [0.594, 1.010] | 3.37 [2.30, 5.74] | 1 | 2.70 [2.20, 3.59] |
| Boxcar | 0.488 [0.367, 0.691] | 6.36 [3.83, 11.13] | 5<sup>*</sup> | 3.11 [2.45, 4.43] |
| *Scenario C: recovery intervals only (ℓ<sub>R</sub>)* |  |  |  |  |
| GLCT | – | 6.42 [6.04, 6.86] | 4 | – |
| Geometric | – | 6.39 [5.36, 7.74] | 1 | – |

**Figure 1:**
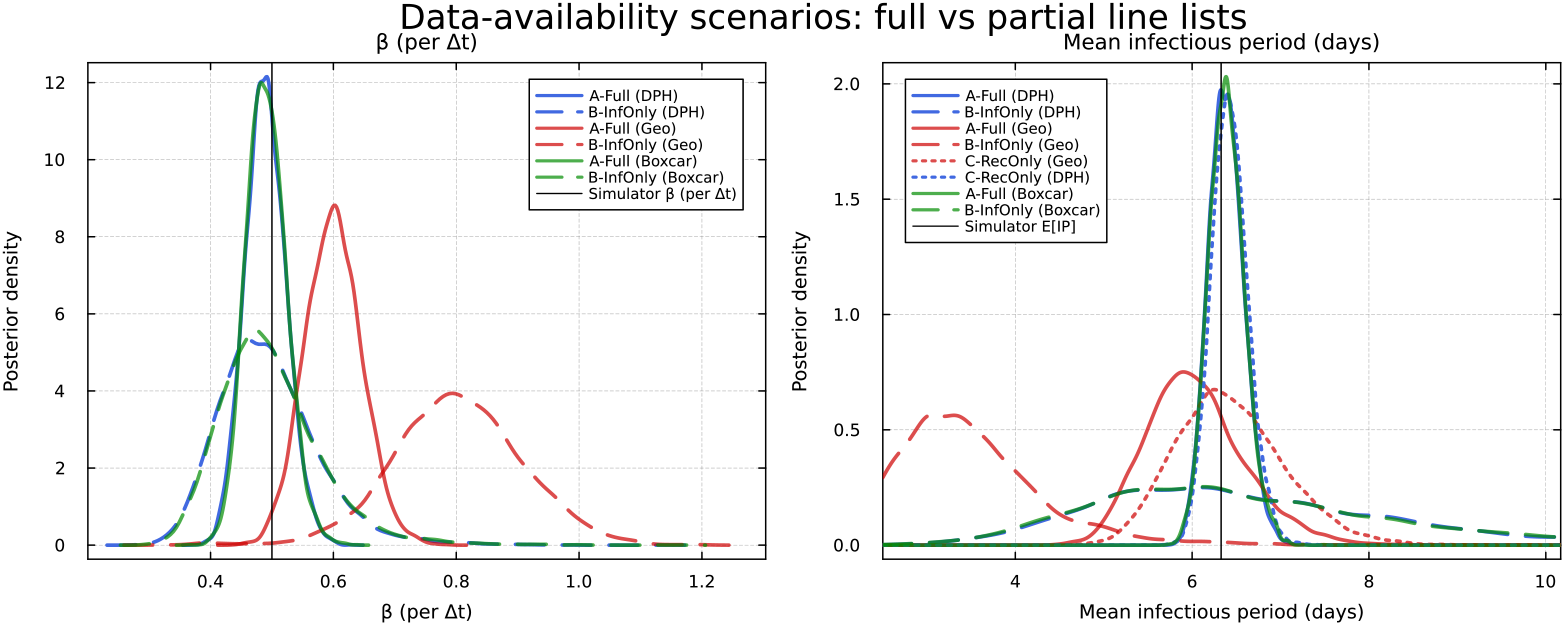
Posterior marginal densities in the synthetic data-availability study. Left: transmission rate *β* (per step, Δ*t* = 1 d) under Scenarios A (full line list; solid) and B (infection times only; dashed). Right: mean infectious period (days) under Scenarios A, B and C (recovery intervals only; dotted). Colours denote the infectious-period model: NegBin DPH via GLCT (blue), geometric (red) and Boxcar (green). Vertical lines mark the values of the generating discrete map (*β* = 0.5, 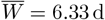).

**Figure 2:**
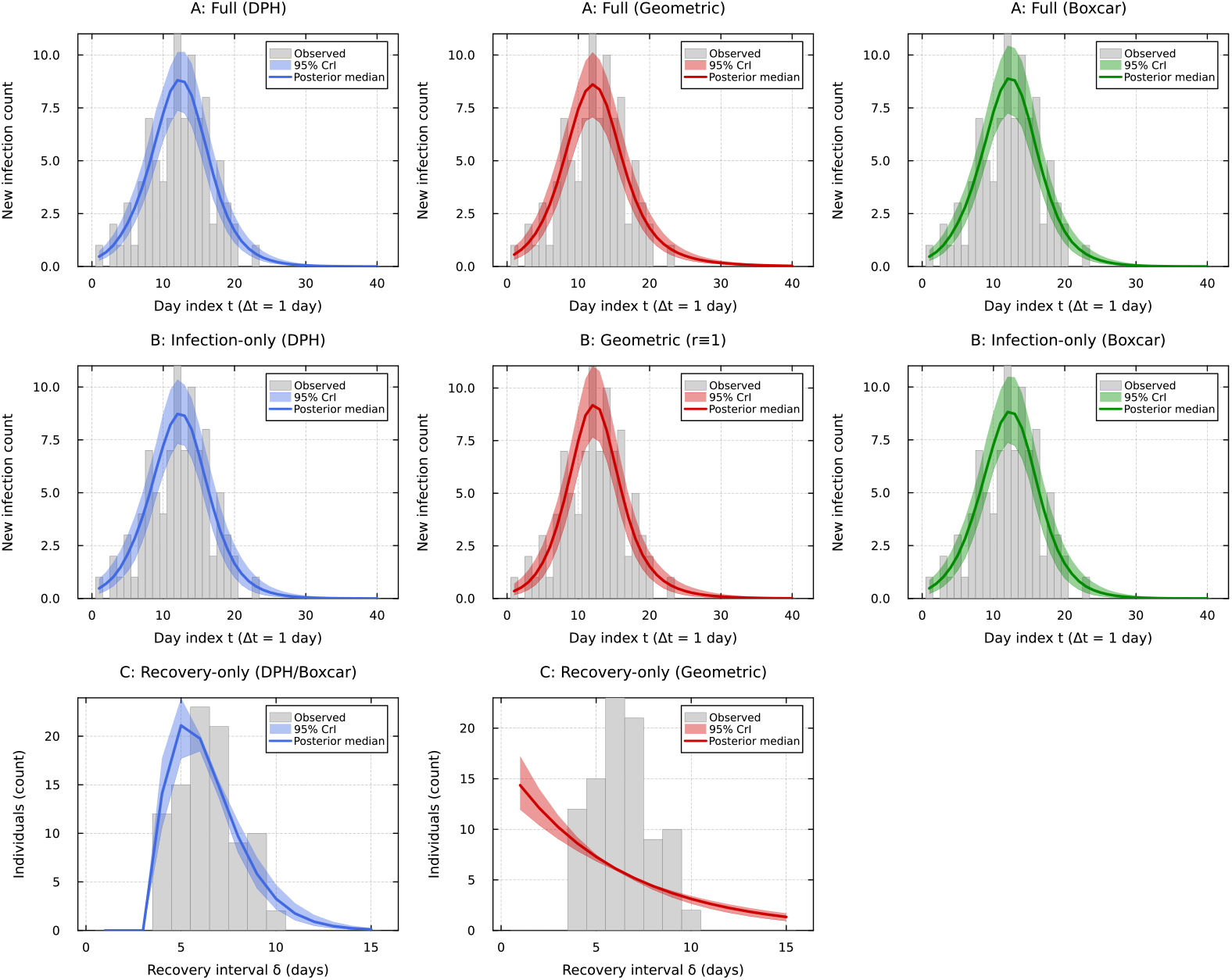
Posterior predictive checks. Columns: infectious-period model (NegBin DPH via GLCT, geometric, Boxcar). Top and middle rows: epidemic curves for Scenarios A and B; each panel shows observed daily incidence (grey bars) with the posterior median (solid line) and 95% credible band (shaded) of the expected daily incidence (parameter uncertainty only). Bottom row: Scenario C, observed recovery intervals (grey bars) against the expected counts, from 1 d, under the fitted NegBin DPH (left; identical for the Boxcar model) and geometric (centre) recovery distributions.

In the model using the geometric distribution (*r* ≡ 1), the misspecification did not show up in the infection-time posterior predictive checks (Figure 2). However, it biased the estimates depending on which data were available. With the full line list, the recovery intervals held the mean infectious period near its true value, so the wrong shape was absorbed by *β* and 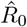 was inflated by 14%. In the case of infection times only, *R*_0_ was held by the final size and stayed close to the value implied by the observed attack rate of 0.92 under the final-size relation (*R*_0_ ≈ 2.74), with an interval that contained the true value. The misspecification appeared instead in *β*, inflated by 60%, and in the mean infectious period, shortened by 47%. The same bias direction appeared in outbreaks simulated from a stochastic chain-binomial model (Supplementary Section S4).

### 3(b) DDSA vs count-based compartmental inference

This analysis explores the benefits of recording individual recovery times in addition to incidence. DDSA was compared with a continuous-time compartmental ODE fitted to aggregated daily incidence counts under Poisson or Negative-Binomial observation models [43], on the synthetic outbreak of Section 3(a). The count model was fitted using four different configurations: an exponential infectious period with a Poisson likelihood, an exponential infectious period with a Negative-Binomial likelihood, a staged Erlang-4 infectious period with a Negative-Binomial likelihood, and an exponential infectious period with the removal rate fixed at 1*/µ* (where *µ* = 4 d is the mean of the continuous-time Erlang-4 infectious period from which the synthetic DPH is built). The count models relied only on daily incidence counts. The first three models estimated the removal rate jointly with the transmission rate. In contrast, DDSA used the complete line list information, which included the infection day and recovery interval for each case, along with the final count of cases out of the 99 individuals at risk. All summaries were compared on the calendar-day scale.

With the removal rate estimated, the three count models gave posterior medians for *R*_0_ of 4.6–7.5 (truth 3.16), with 95% credible intervals within [1.76, 20.16], and mean infectious periods with 95% credible intervals within [1.7, 47.2] d. This ambiguity persisted regardless of whether the shape of the infectious period was correctly staged (Figure 3). Fixing the mean infectious period at 4 d narrowed the *R*_0_ posterior but biased it below the truth (*R*_0_ = 2.75 [2.36, 3.15]), because the assumed rate overestimated removal relative to the effective discrete infectious period of 6.33 d.

**Figure 3:**
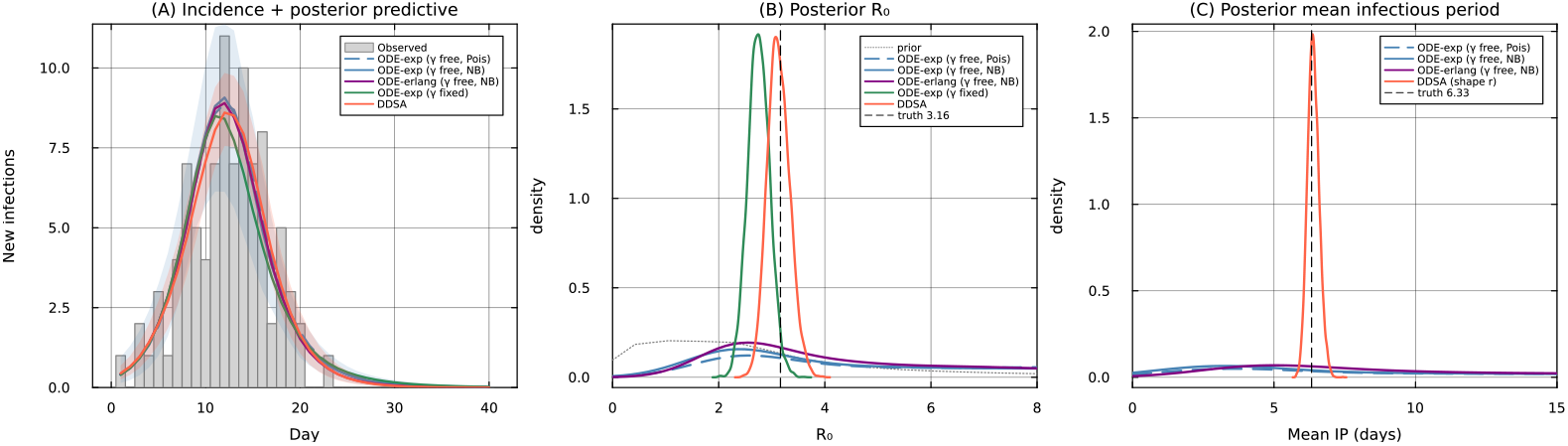
DDSA versus count-based compartmental inference on one synthetic Erlang-4 outbreak (true *R*_0_ = 3.16, mean infectious period 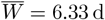. (A) Observed daily incidence with posterior medians of the expected daily incidence for all five models, and 95% credible bands for the exponential Negative-Binomial count model and DDSA (parameter uncertainty only). (B) Posterior *R*_0_ for the count-ODE fits with a free removal rate (exponential, Erlang), the fit with the mean infectious period fixed at 4 d (green) and DDSA (orange), with the prior (dotted) and true value (dashed). (C) Posterior mean infectious period for the fits with a free removal rate and DDSA. All five models used four chains, with 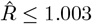, bulk ESS ≥ 1,133 and no divergent transitions.

DDSA, which also uses the recovery intervals and final size, recovered *β*, the mean infectious period and *R*_0_ (Table 4, Scenario A, GLCT). Every draw used *r* = 4, the true value and the largest allowed by the shortest recorded interval of 4 d, so this concentration reflects both the cap and the data (Supplementary Section S2.3). All five models reproduced the observed incidence curve (Figure 3A), so they differed in what they identified.

Adjusting one factor at a time (see Supplementary Section S2) showed that the loss of information on *R*_0_ in the count models during this outbreak with a high attack rate (91 out of 99 infected individuals; the loss was smaller at moderate attack rates) can be attributed to a single likelihood term. Specifically, the Poisson and Negative Binomial count models do not include the never-infected term (*N* − *K*) log(1 − *τ*) found in the Binomial final size equation.

A count model with a never-infected cell recovered the DDSA estimate of *R*_0_ from infection times alone (Scenario B), with either the discrete map or an ODE. The incidence curve fixes the early growth rate, along which *R*_0_ and the mean infectious period trade off [26, 44], and the recovery intervals, which counts do not contain, fix the position along this ridge. In one simulated outbreak at each size up to 10,000, the count models moved towards the truth as the population grew, with the discrete map and an ODE giving similar *R*_0_. However, the never-infected count gave DDSA much narrower credible intervals across all population sizes (as detailed in Supplementary Section S2). Table 5 summarises what each data type identifies.

**Table 5:** What each data type identifies (synthetic Erlang-4 outbreak, *N* = 99). “Yes”: the posterior concentrates near the true value; “weak”: identified only within a wide interval; “no”: not identified by the data.

| **Data used** | **Likelihood terms** | **Growth rate** | *R*<sub>0</sub> | **Mean IP** | **IP shape** |
|---|---|---|---|---|---|
| Full line list | $\ell_I + \ell_R + \ell_{\text{constr}}$ | yes | yes | yes | yes |
| Infection times + final size | $\ell_I + \ell_{\text{constr}}$ | yes | yes<sup>*a*</sup> | weak | no |
| Counts + never-infected count | Multinomial with a never-infected cell | yes | yes<sup>*a*</sup> | weak | no |
| Daily counts only | Poisson or Negative Binomial | yes | no<sup>*b*</sup> | no<sup>*b*</sup> | no |
| Recovery intervals only | $\ell_R$ | no | no | yes | yes |

<sup>*a*</sup> With few individuals left uninfected (here *N* − *K* = 8) the upper limit depends on the *R*<sub>0</sub> prior bound (Supplementary S2).
<sup>*b*</sup> Bounded only by the priors: the posterior runs along the growth-rate ridge to a prior edge.

### 3(c) Influenza A(H1N1), Washington State University, 2009

DDSA was applied to the line list from the 2009 H1N1 outbreak at Washington State University, the dataset analysed by KhudaBukhsh et al. [12], to compare the two approaches on the same data (*K* = 2,276 infections over 105 days, days 0–104). Recovery intervals, recorded in whole days (Section 2(c)(ii)), entered the likelihood through the geometric PMF of Eq. (2.11), and the 13 recoveries after day 104 were treated as right-censored, following their continuous-time framework. The observation window begins mid-epidemic, implying a substantial initial infected fraction *ρ*, and the population size *N* at risk is unknown. *N* enters only through the final-size constraint, where it is confounded with the attack rate *τ* (*β, γ, ρ*). Priors were therefore placed on *γ* (centred on a mean infectious period of 5.5 d) and on *N* (bounded by the WSU student body [34]; Supplementary Table S1), and all four parameters (*N* , *β, γ, ρ*) were estimated jointly. Because the recovery intervals were generated under an assumed recovery rate, the posterior for *γ* is a consistency check that the DDSA geometric likelihood reproduces them, not an independent estimate of the H1N1 infectious period.

The fitted mean infectious period (5.12 days, 95% CrI [4.94, 5.32]; Table 6) reproduced the mean recorded recovery interval in the line list (5.06 days), and posterior predictive replicates of the final size covered the observed cases. KhudaBukhsh et al. [12] conditioned their likelihood on *n* (initially susceptible population) and imputed it at each iteration of a hybrid Gibbs sampler as *n* ∼ NegBinom(*K, τ*), with *τ* given by the deterministic final-size equation. They also fixed the infectious period at *γ*^−1^ = 5.5 days, whereas DDSA estimated it from the recovery intervals. The DDSA estimate of the effective population size (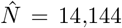; 90% CrI [10,898, 17,453]) was larger than theirs (7,051; 90% CrI [6,602, 7,581]), with a smaller initial infected fraction 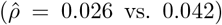, which is confounded with *N* . However, using the same continuous-time likelihood and priors, but replacing their imputation step by the exact conditional draw under the complete Binomial final-size term, or summing *N* out of the same likelihood, moved the estimates of *N* and *R*_0_ towards the DDSA estimates (Supplementary Table S11). The transmission rates agreed (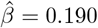 (90% CrI [0.181, 0.200]) vs. 0.189 (90% CrI [0.185, 0.196])). The DDSA reproduction number was slightly lower (0.97 (90% CrI [0.94, 1.02]) vs. 1.06 (90% CrI [1.04, 1.09])), consistent with the larger inferred *N* , which implies a smaller attack rate, and both estimates are close to 1. The fitted curve followed the overall decline in incidence but not its two early peaks (days 10 and 17; Figure 4A), as both analyses use a deterministic mean-field SIR model and assume a constant transmission rate, which produces a single, smooth epidemic curve. The posterior summaries can be found in Table 6, and the marginals are illustrated in Figure 4.

**Table 6:**
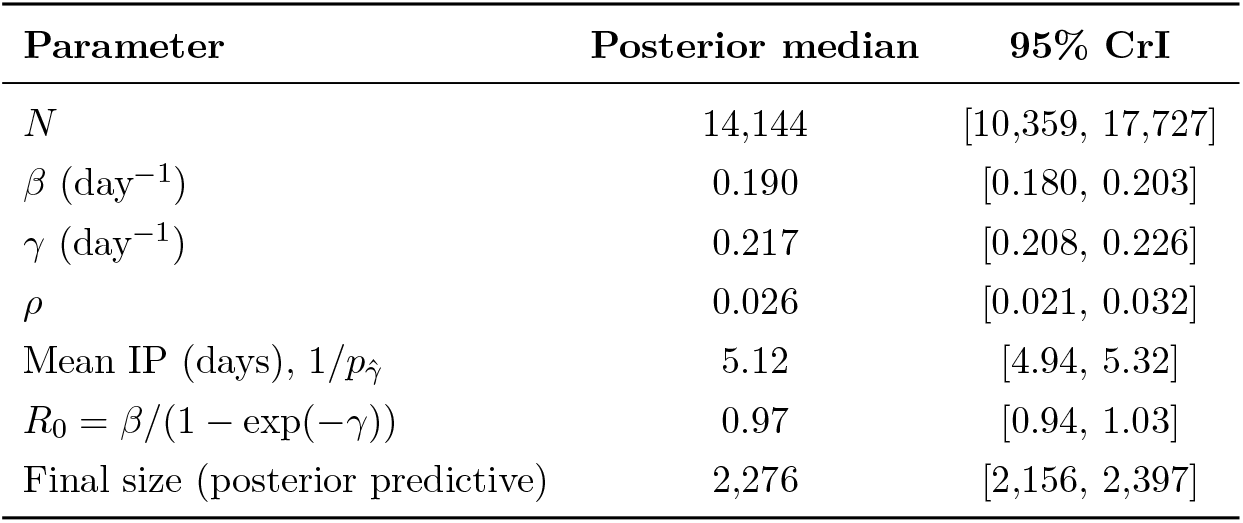
Posterior summary for the DDSA fit to the 2009 H1N1 outbreak at Washington State University. All four parameters (*N, β, γ, ρ*) are inferred jointly; derived quantities (Mean IP, *R*_0_, final size) are computed sample-by-sample from the chain (the final size as a replicate *K* ∼ Binomial(*N, τ*)). Convergence: 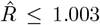and bulk ESS ≥ 2,974 across all parameters including *N* . Observed final size *K* = 2,276.

| **Parameter** | **Posterior median** | **95% CrI** |
|---|---|---|
| *N* | 14,144 | [10,359, 17,727] |
| β (day<sup>−1</sup>) | 0.190 | [0.180, 0.203] |
| γ (day<sup>−1</sup>) | 0.217 | [0.208, 0.226] |
| ρ | 0.026 | [0.021, 0.032] |
| Mean IP (days), 1/*p*<sub>γ̂</sub> | 5.12 | [4.94, 5.32] |
| *R*<sub>0</sub> = β/(1 − exp(−γ)) | 0.97 | [0.94, 1.03] |
| Final size (posterior predictive) | 2,276 | [2,156, 2,397] |

**Figure 4:**
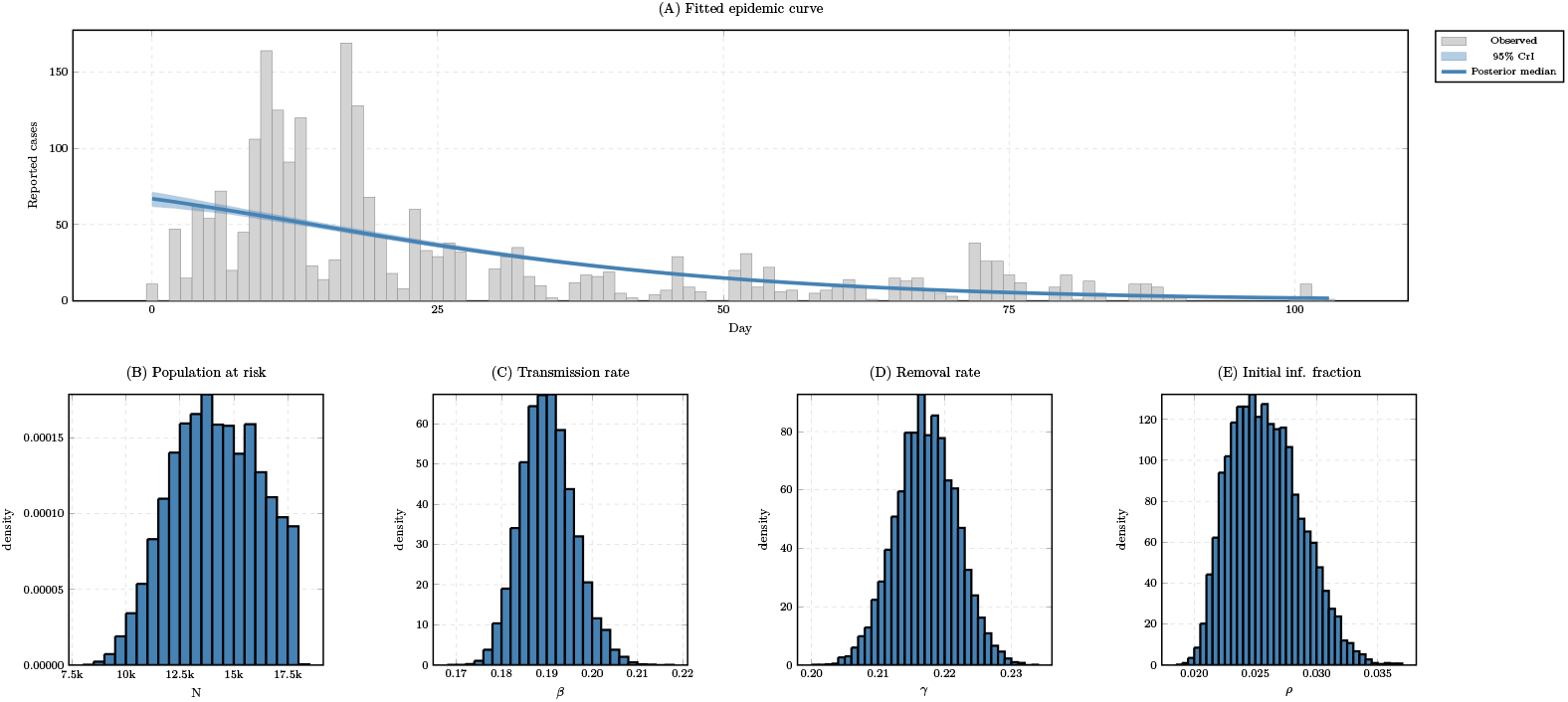
DDSA fit to the 2009 H1N1 outbreak at Washington State University (*K* = 2,276 cases over 105 days). Fitted epidemic curve: posterior median and 95% credible band of the expected daily incidence (parameter uncertainty only), overlaid on the observed daily incidence. Posterior marginals for the population at risk *N* , *β, γ*, and *ρ* are shown alongside.

### 3(d) Ebola virus disease, Democratic Republic of the Congo, 2018

The 2018 Ebola outbreak in the Democratic Republic of the Congo comprised three epidemiologically distinct waves, which were analysed separately using the base discrete SIR model with geometric infectious periods and a wave-specific effective population size *N* (Table 7, Figure 5). Within each wave, records were partitioned by data completeness. Complete records (onset and hospitalisation both observed) contributed *ℓ*_*I*_ + *ℓ*_*R*_ + *ℓ*_constr_, with hospitalisation as the removal-time proxy (**δ** = hosp − onset). Onset-only records contributed *ℓ*_*I*_ + *ℓ*_constr_. The few hospitalisation-only records entered through a convolution over latent onset *u*:

**Table 7:**
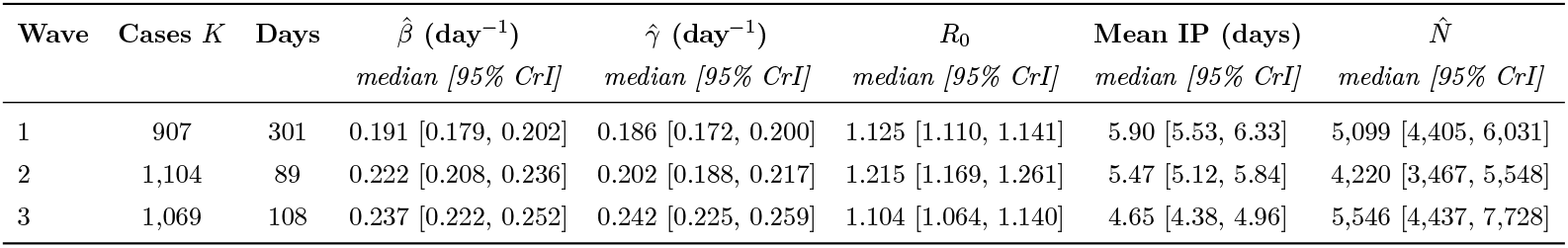
Posterior medians and 95% credible intervals for the three independently analysed waves of the 2018 Ebola outbreak in the Democratic Republic of the Congo, fitted with the discrete SIR model and all four parameters (*β, γ, ρ, N*) inferred jointly.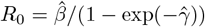 and mean 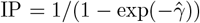 are derived quantities of the fitted discrete-time process, computed sample-by-sample from the chain. Convergence: across all parameters and waves, max 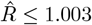, min bulk ESS ≥ 1,700, and no divergent transitions (out of 8,000 draws per wave).

| **Wave** | **Cases *K*** | **Days** | **β̂ (day<sup>−1</sup>)** | **γ̂ (day<sup>−1</sup>)** | *R*<sub>0</sub> | **Mean IP (days)** | *N̂* |
|---|---|---|---|---|---|---|---|
|  |  |  | *median [95% CrI]* | *median [95% CrI]* | *median [95% CrI]* | *median [95% CrI]* | *median [95% CrI]* |
| 1 | 907 | 301 | 0.191 [0.179, 0.202] | 0.186 [0.172, 0.200] | 1.125 [1.110, 1.141] | 5.90 [5.53, 6.33] | 5,099 [4,405, 6,031] |
| 2 | 1,104 | 89 | 0.222 [0.208, 0.236] | 0.202 [0.188, 0.217] | 1.215 [1.169, 1.261] | 5.47 [5.12, 5.84] | 4,220 [3,467, 5,548] |
| 3 | 1,069 | 108 | 0.237 [0.222, 0.252] | 0.242 [0.225, 0.259] | 1.104 [1.064, 1.140] | 4.65 [4.38, 4.96] | 5,546 [4,437, 7,728] |

**Figure 5:**
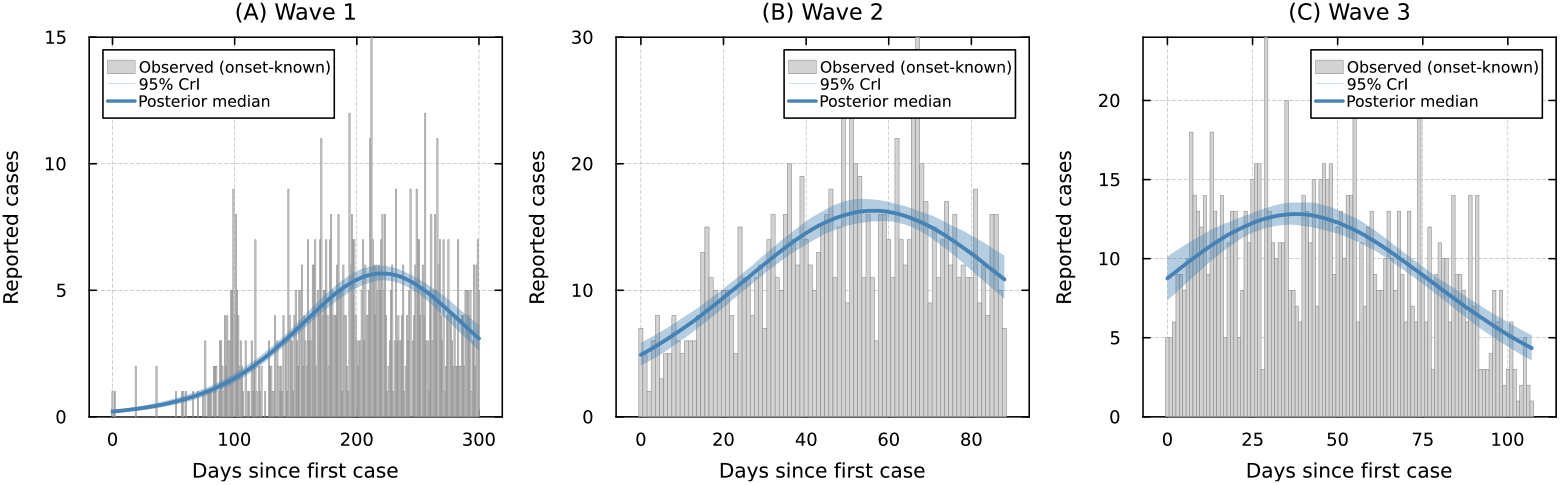
DDSA applied to the three waves of the 2018 Ebola virus disease outbreak in the Democratic Republic of the Congo. Each wave was analysed independently with *N* inferred as a free parameter. Panels show the posterior median and 95% credible band of the expected daily number of onsets (parameter uncertainty only), overlaid on the observed daily onsets of records with a known onset date, for Waves 1–3.

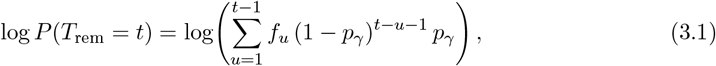

summing over all feasible onset days *u < t* under the same geometric infectious-period distribution.

Posterior median *R*_0_ exceeded one in every wave, conditional on hospitalisation ending transmission, and was highest in Wave 2 (Table 7), the same pattern Vossler et al. [14] reported for their continuous-time *β/γ* (1.124, 1.214, 1.098). The fitted mean infectious period shortened from Wave 1 to Wave 3 as the per-day removal probability 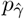 rose by about 27%, the same rise as in Vossler et al. The lower *R*_0_ in Wave 3 than in Wave 2 was therefore associated with faster removal of cases, that is, a shorter interval from onset to hospitalisation, rather than with a fall in the transmission rate *β*. This pattern is also consistent with their reported intensified quarantine and isolation in treatment centres. However, within this model, faster removal cannot be distinguished from faster case finding. The estimates of *β* and *ρ* also agreed with theirs within sampling error in all three waves, and 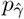 closely matched their continuous-time *γ* (Wave 3: 0.215 [0.201, 0.228] against 0.214 [0.199, 0.230] d^−1^). In addition, Roy et al. [15] analysed the same wave from daily incidence alone, with a count-based DSA likelihood on a Poisson contact network. Their *R*_0_ = 1.121 [1.044, 1.200] agreed with the DDSA estimate (1.104 [1.064, 1.140]) and with that of Vossler et al. (1.098 [1.061, 1.135]). Their rate 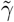, which in their model combines removal and per-contact transmission, was of similar size to the removal rates above, with a wider interval (0.205 [0.134, 0.426] d^−1^). They attribute this to making no assumption about exact event times. This agrees with the results of Section 3(b) and Supplementary Section S2: incidence constrains the infectious period through the epidemic curve, whereas individual onset-to-hospitalisation intervals constrain it directly.

Vossler et al. [14] estimated one overall effective population size, *N* ≈ 16,000, for the entire outbreak, whereas a separate 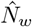 was fitted here for each wave. Each wave has its own final-size term, *K*_*w*_ ∼ Binomial(*N*_*w*_, *τ*_*w*_), and the outbreak moved between health zones from wave to wave [14]. Fitting the waves separately prevents them from sharing information about *N* . The three 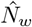 are not additive, because Waves 1 and 3 are centred on the same northern health zones and their at-risk populations likely overlap. Nevertheless, both analyses inferred an effective susceptible pool far smaller than the demographic totals, consistent with the impact of ring vaccination, contact tracing, and quarantine.

### 3(e) Measles, Hagelloch, Germany, 1861

The 1861 measles outbreak in Hagelloch, Germany, comprised *K* = 188 cases over 86 days [24]. Infectious-period durations were computed as **δ**_*i*_ = (rash date − prodrome date) + 4 days, reflecting the assumption of Neal and Roberts [36] that children were infectious from one day before the prodrome until three days after the rash appeared. These durations had a mean of 7.95 days and a coefficient of variation of 0.22, below that of a geometric distribution with the same mean (0.94), so the infectious period was modelled with a Negative Binomial DPH. Since every duration is at least **δ**_min_ = 4 days, an offset Negative Binomial DPH was used. This model consisted of **δ**_min_ − *r* deterministic one-day stages followed by *r* geometric stages with progression probability *p*. Its support starts at **δ**_min_, its mean is 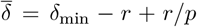, and it requires that *r* ≤ **δ**_min_. The model sampled 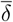 and set 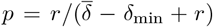, so that 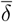 keeps its meaning across values of *r*. It was assumed that a single index case from Class 2 initiated the outbreak, so the initial infectious fraction was fixed at *ρ* = 1*/N*_2_ for Class 2 and zero for Classes 0 and 1.

The line list partitions the population into three classes: pre-school children (Class 0, *K*_0_ = 90), younger school students (Class 1, *K*_1_ = 30), and older school students (Class 2, *K*_2_ = 68, including the index case). Between-group transmission was captured using a 3 *×* 3 who-acquires-infection-from-whom (WAIFW) matrix **B**, where *B*_*ij*_ is the per-day transmission rate to group *i* from group *j*:

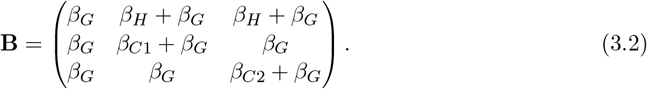

The per-step force of infection on group *i* is then 1 − exp(− Λ_*i*_), where Λ_*i*_ = ∑_*j*_ *B*_*ij*_ *I*_*j*_ and *I*_*j*_ denotes the infectious fraction of group *j*. Within-class transmission is governed by *β*_*C*1_ (Class 1) and *β*_*C*2_ (Class 2), a global term *β*_*G*_ acts across all groups, and a school-to-preschool term *β*_*H*_ captures transmission from infectious school students to pre-school children. This structure is a mean-field, group-level approximation of the individual-level contact model of Neal and Roberts [36] in its homogeneous-mixing form (their M_*θ*_ model without distance decay). The rates *β*_*C*1_, *β*_*C*2_ and *β*_*G*_ correspond to their classroom and global routes, and *β*_*H*_ acts as a proxy for their same-household term. In their model, the household term is applied between any two members of the same household. However, as the compartmental model does not represent this, a single household rate was restricted to the school-to-preschool direction. This assumption was based on the pre-school group forming a secondary wave in the line list, in which most cases were attributed to school-age children (53 of 90), 41 of them within the same family [24]. Preschool-to-school and within-preschool household transmission therefore enter through *β*_*G*_.

The GLCT map was the primary analysis, as it represents the same Negative Binomial family with a smaller state vector than the Boxcar map, which was fitted as an implementation cross-check. The integer-valued parameters (*r, N*_0_, *N*_1_, *N*_2_) were summed out of the likelihood. The class sizes were inferred through Binomial final-size constraints, with prior ranges bounded by the 197 children under 14 recorded by Pfeilsticker [24] (Supplementary Table S3). Neal and Roberts [36] instead fixed the population sizes to match the case count (188 individuals, all infected), which assumes a 100% attack rate.

Posterior summaries are shown in Table 8 and Figures 6–7. The global *R*_0_ agreed between the two maps (2.17 under both). The phase count reached its upper limit, with *r* = min_*i*_ **δ**_*i*_ = 4, in nearly every draw, indicating that the cap was binding. At *r* = 4, the recovery-interval likelihood was still increasing, and the estimated infectious period showed more variability than the recorded durations (Supplementary Section S4.3). The Boxcar fit used the same law and has the same cap, so the agreement between the two fits checks the implementation rather than the law. A binding cap can bias *R*_0_ upward (Supplementary Section S4). The fit gave *β*_*C*1_ *> β*_*C*2_ with *P* (*β*_*C*1_ *> β*_*C*2_) = 0.996, in qualitative agreement with Neal and Roberts [36], who found strong evidence of transmission within classroom 1 and weaker evidence of transmission within classroom 2. For Class 1, the model-implied attack rate closely matched the observed one (Table 8), whereas attack rates for Classes 0 and 2 were underpredicted by about 15%. The fitted epidemics also peaked closer together in time than observed. The model peaked on day 26 in Class 1 and on day 28 in Classes 0 and 2, whereas the largest daily counts fell on days 22, 35 and 32 in Classes 1, 0 and 2 (Figure 7). These differences were consistent with the mean-field approximation, which averages over children with heterogeneous, localised household exposures. Finally, the posteriors for *N*_0_ and *N*_2_ concentrated near their upper bounds, and the total across the three classes, about 210, exceeded the number of children who could have been infected. This may reflect the smaller final size of the mean-field model rather than a larger population at risk. Fixing the class sizes at the case counts gave a similar *R*_0_ (2.17) and the same ordering *β*_*C*1_ *> β*_*C*2_.

**Table 8:**
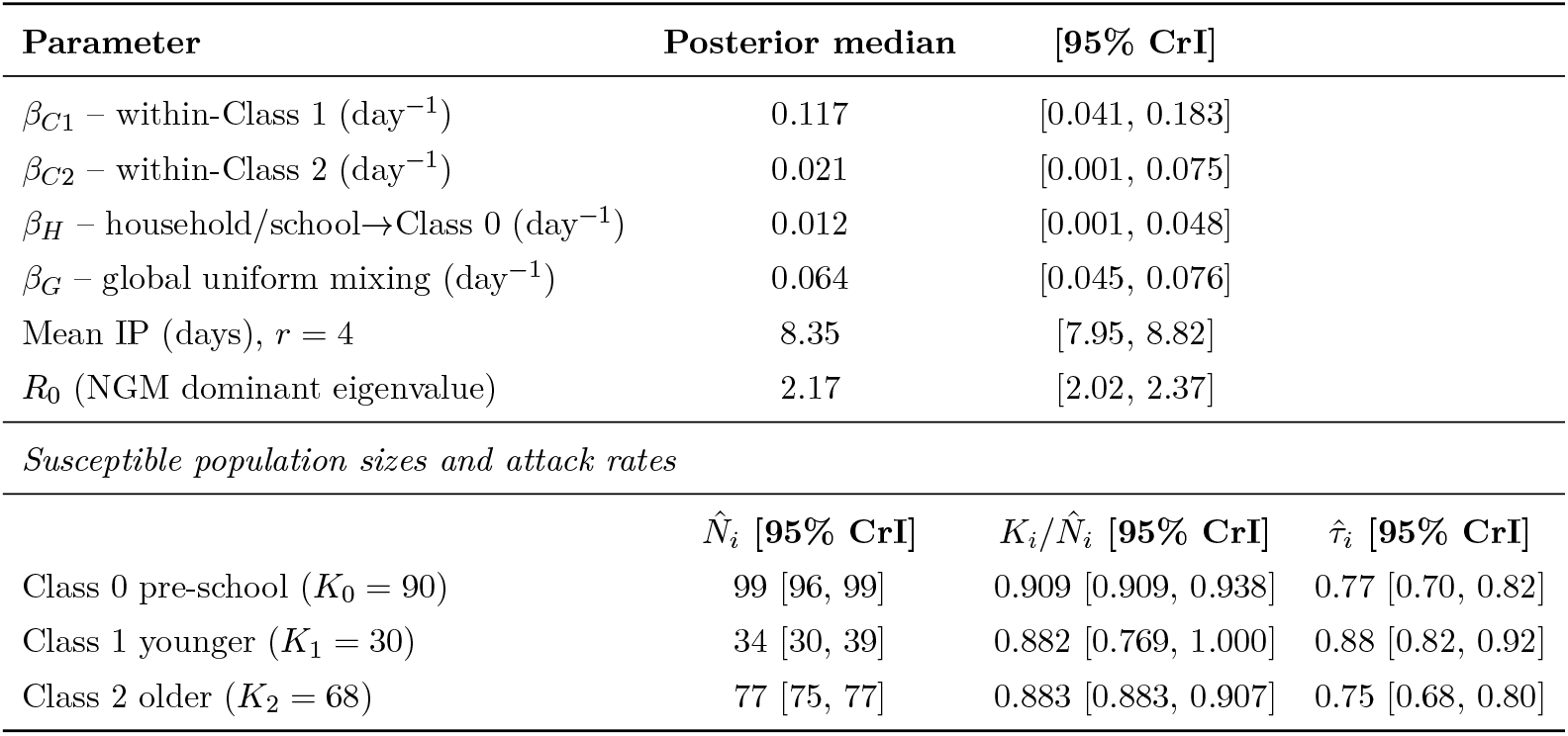
Posterior medians and 95% credible intervals for the three-group M_*θ*_-analogue DDSA (GLCT and Boxcar maps, NegBin DPH) applied to the Hagelloch 1861 measles outbreak. 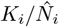 is the observed attack rate of class *i* and 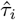 the model-implied attack rate predicted by the fitted dynamics; *R*_0_ is the dominant eigenvalue of the 3 *×* 3 next-generation matrix. All derived quantities are computed sample-by-sample from the chain. Convergence: all parameters reach min. ESS_bulk_ ≥ 1,551 and max.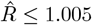 under both maps. *r* = 4 in every GLCT draw and in 7,999 of 8,000 Boxcar draws.

| **Parameter** | **Posterior median** | **[95% CrI]** |  |
|---|---|---|---|
| β<sub>C1</sub> – within-Class 1 (day<sup>−1</sup>) | 0.117 | [0.041, 0.183] |  |
| β<sub>C2</sub> – within-Class 2 (day<sup>−1</sup>) | 0.021 | [0.001, 0.075] |  |
| β<sub>H</sub> – household/school→Class 0 (day<sup>−1</sup>) | 0.012 | [0.001, 0.048] |  |
| β<sub>G</sub> – global uniform mixing (day<sup>−1</sup>) | 0.064 | [0.045, 0.076] |  |
| Mean IP (days), *r* = 4 | 8.35 | [7.95, 8.82] |  |
| *R*<sub>0</sub> (NGM dominant eigenvalue) | 2.17 | [2.02, 2.37] |  |
| *Susceptible population sizes and attack rates* |  |  |  |
|  | *N̂*<sub>*i*</sub> **[95% CrI]** | *K*<sub>*i*</sub>/*N̂*<sub>*i*</sub> **[95% CrI]** | *τ̂*<sub>*i*</sub> **[95% CrI]** |
| Class 0 pre-school (*K*<sub>0</sub> = 90) | 99 [96, 99] | 0.909 [0.909, 0.938] | 0.77 [0.70, 0.82] |
| Class 1 younger (*K*<sub>1</sub> = 30) | 34 [30, 39] | 0.882 [0.769, 1.000] | 0.88 [0.82, 0.92] |
| Class 2 older (*K*<sub>2</sub> = 68) | 77 [75, 77] | 0.883 [0.883, 0.907] | 0.75 [0.68, 0.80] |

**Figure 6:**
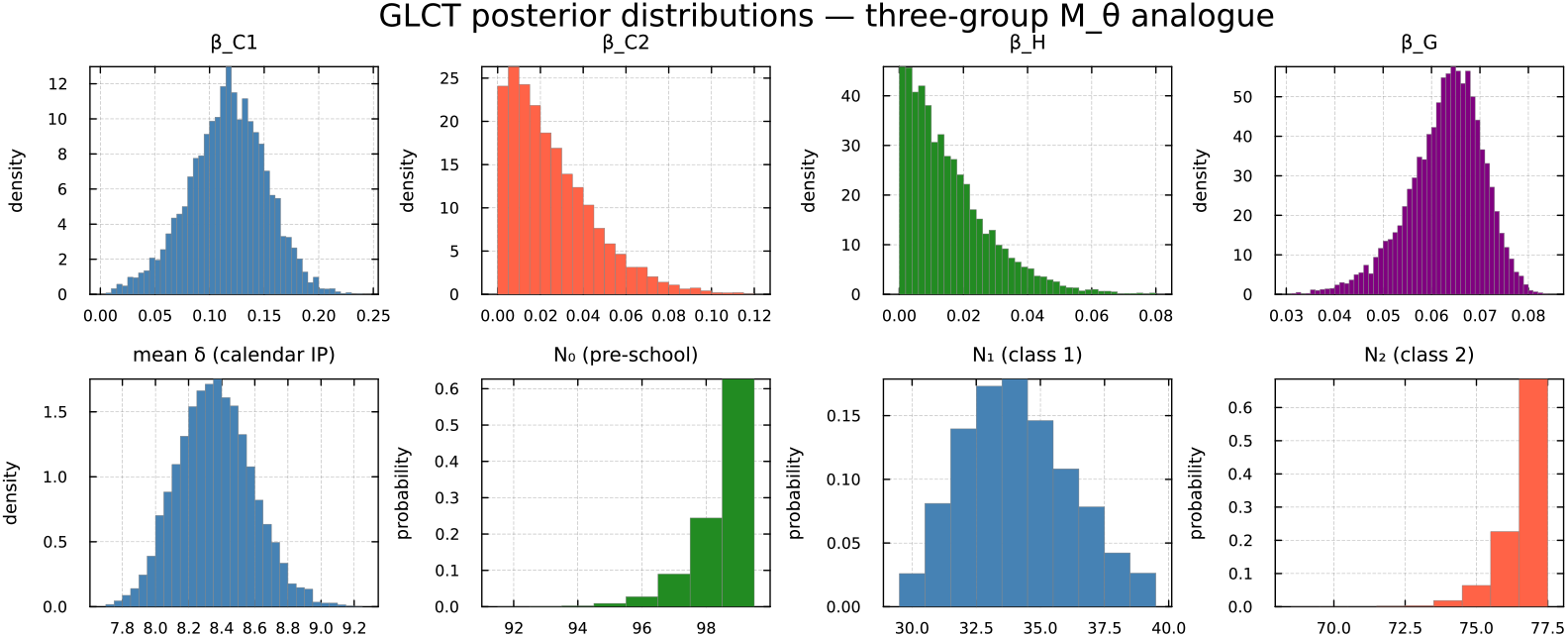
Posterior marginal distributions for the three-group M_*θ*_-analogue DDSA applied to the Hagelloch 1861 measles outbreak. GLCT fit. Top row: transmission rates *β*_*C*1_, *β*_*C*2_, *β*_*H*_ and *β*_*G*_. Bottom row: mean infectious period (calendar days) and the inferred susceptible population sizes *N*_0_, *N*_1_, *N*_2_, shown as posterior probabilities (one bar per value).

**Figure 7:**
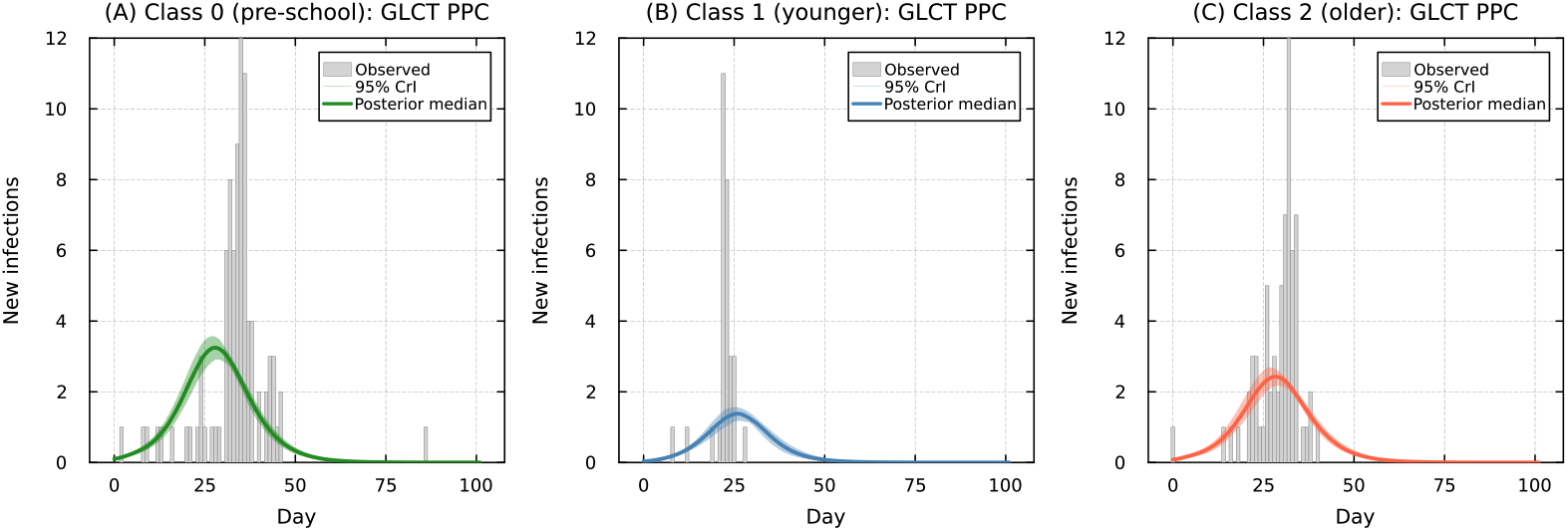
Fitted epidemic curves for the three-group M_*θ*_-analogue DDSA (GLCT fit), Hagelloch 1861 measles outbreak. Each panel shows observed daily infections (bars) with the posterior median (solid line) and 95% credible band (shaded) of the expected daily infections in that class, which sum to the model’s predicted number of cases. The band reflects parameter uncertainty propagated through the deterministic recurrence map.

## 4 Discussion

DDSA offers a method to estimate key epidemiological parameters, including the reproduction number, the distribution of the infectious period, and the effective size of the population at risk, directly from line lists. It reformulates the survival likelihood of dynamic survival analysis [12, 13] in discrete time, so that individual infection times, infectious-period durations and final size are evaluated on the calendar-day grid of the line list. DDSA has three practical advantages. (i) No continuous solution has to be interpolated onto observed dates. (ii) The infection-time PMF and the discrete phase-type recovery distribution are computed exactly by iterating the recurrence map. As a result, a non-geometric infectious period requires neither a numerical PDE solve nor a numerical convolution. (iii) The likelihood can therefore be written directly in a standard probabilistic programming language and sampled with off-the-shelf NUTS, without model-specific numerical code. Continuous-time DSA can also be sampled with NUTS, as our re-implementations were (Supplementary Section S3), but only with the addition of ODE and PDE solvers, which the discrete maps replace. DDSA’s estimates are comparable to those of continuous-time DSA. On simulated outbreaks, the two methods agreed except when the daily step was coarse relative to epidemic growth (Supplementary Section S3; Limitations). The continuous-time re-implementations used for this comparison were checked against the published codes [12, 13]. Computational speed was not assessed in this study, and a comparison of sampling efficiency (ESS per second) with continuous-time DSA is reserved for future work.

Synthetic experiments based on one outbreak of 99 people, with checks at larger population sizes (Supplementary Section S2), suggest that the infection-time and recovery components of the likelihood play different inferential roles (Table 5). Infection times determine the early growth rate, along which *R*_0_ and the mean infectious period trade off [26, 44]. When the at-risk population is known, the number of never-infected individuals bounds *R*_0_. Recovery intervals identify the infectious period, which, together with the growth rate, fixes *R*_0_. Its shape is identified within the one-stage-per-day NegBin DPH used here, whose number of stages cannot exceed the shortest recorded interval (Limitations). These roles also determine where a mis-specified infectious period becomes visible. Under a geometric infectious period, the incidence curves remained visually similar to those of the correctly specified DPH models. Nevertheless, the estimates were biased in a direction set by the available data. With the full line list, the recovery intervals held the mean infectious period near its true value, and *R*_0_ was inflated by 14%. With infection times only, the final size held *R*_0_ near the value implied by the observed attack rate (posterior median 2.70, against 2.74 from the final-size relation; truth 3.16, inside the 95% CrI), while *β* was inflated by 60% and the mean infectious period was roughly halved. Only the recovery intervals revealed the misspecification. The geometric fit missed their peak at 6–7 d and placed about 37 of the 92 intervals at 1–3 d, where none were observed (Figure 2, bottom row). In this example, therefore, checks of the incidence curve did not detect a misspecified infectious-period shape, whereas a check of the recovery intervals did.

Comparing DDSA with count models separates what is lost by aggregating the data from what is lost by choosing a count likelihood. In this high-attack-rate outbreak, Poisson and Negative-Binomial count models did not identify *R*_0_ because they omit the individuals who were never infected, not because the data were aggregated. With *N* known, a count model with a never-infected cell recovered the DDSA estimate of *R*_0_ from infection times alone, without recovery intervals and without fixing the removal rate. At lower attack rates, a Poisson count model also constrained *R*_0_, although less precisely (Supplementary Section S2), so this loss should not be generalised to all incidence-only analyses. Without the never-infected count, *R*_0_ was constrained closely only once the removal rate was fixed, consistent with KhudaBukhsh et al. [12] and Putter et al. [45], who also fixed the recovery-time distribution because only incidence was observed. The estimate was then only as accurate as the fixed rate. Here, an assumed mean of 4 d biased *R*_0_ below the truth. Aggregation does matter for the infectious period. Without recovery intervals, its shape is not identified, and its mean is identified only weakly, through *R*_0_ and the growth rate. This follows from data resolution rather than from the likelihood formulation. Roy et al. [15] developed a marginal likelihood for count data that treats summarised counts as interval-censored survival observations in continuous time. When the population at risk is known, our count model with a never-infected cell is equivalent to their likelihood (Supplementary Section S2). The two approaches are complementary. Count-based DSA needs only incidence data, which are more widely available, whereas line lists add individual recovery intervals, which identify the infectious period directly. Where data collection can be planned, these results suggest recording, in addition to onset dates, the date each case stops being infectious (or a removal proxy such as hospitalisation) and the size of the population at risk.

Surveillance line lists are rarely complete. In the analyses considered here, the DDSA likelihood served as the core observation model. It was supplemented with survivor terms for right-censored recoveries in the H1N1 case study, and with a convolution over latent onset times for hospitalisation-only Ebola records. Both are discrete counterparts of terms in the continuous-time DSA likelihood [12, 13]. Records without a recovery or removal date contributed only to the infection-time and final-size terms, as in Scenario B and for the Ebola onset-only records. This allows incomplete line lists to be handled within the same general framework, without ad hoc imputation.

The case studies benchmark the DDSA inference framework against published continuous-time DSA analyses of the same data. They do not test whether an SIR model describes these outbreaks accurately. For H1N1 at WSU, fitting the line list analysed by KhudaBukhsh et al. [12] reproduced the mean recorded recovery interval and gave a transmission rate similar to theirs, with a slightly lower reproduction number and a larger effective population size. How the population size was treated explains much of these differences (Section 3(c); Supplementary Table S11). Across the three Ebola waves, DDSA reproduced the trends of the continuous-time DSA fit of Vossler et al. [14]. The removal rate *γ* rose from Wave 1 to Wave 3, a shortening of onset-to-hospitalisation, while *β* did not fall. Posterior medians suggest *R*_0_ was highest in Wave 2, but this ordering depends on the removal definition and on credible intervals that likely understate uncertainty. These agreements partly reflect shared modelling choices. Both analyses use a mean-field SIR model with the same removal definition, and for H1N1 the recovery times were themselves generated by KhudaBukhsh et al. from the exponential infectious period (mean 5.5 d) assumed in their analysis.

The Hagelloch case study was used to stress-test DDSA on a heterogeneous, group-structured outbreak, where school-centred and within-household transmission depart from homogeneous mixing. The three-group model needed only a 3 *×* 3 transmission matrix in the same recurrence map. It reproduced the qualitative finding of the individual-level analysis of Neal and Roberts [36], with more transmission within classroom 1 than within classroom 2 (*P* (*β*_*C*1_ *> β*_*C*2_) = 0.996). The GLCT and Boxcar implementations gave closely matching summaries (*R*_0_ = 2.17), an internal check that the two discrete representations agree when the dwell-time family is shared. The stress test also exposed the limits of the mean-field approximation in this setting. The model underpredicted the attack rates of Classes 0 and 2, brought the class epidemics closer together in time than observed, and placed *N*_0_ and *N*_2_ near their upper bounds. The stage cap was also binding (Section 3(e)).

More generally, other compartmental structures can be accommodated within DDSA provided the mean-field basis of the survival likelihood is preserved. Additional compartments and further stratification into demographic groups or spatial patches, for example, all follow from the same difference-equation construction, as the Hagelloch model illustrates. These structures assume a closed population in which each individual is infected at most once. As a result, the susceptible population *S*_*k*_ only decreases, and the final-size term applies. Extending DDSA to endemic transmission, with births, waning immunity or reinfection, would require changes to the map and to the likelihood. The map would need inflows to the susceptible class and initial conditions near the endemic state rather than a small seed. Additionally, since *S*_*k*_ would no longer only decrease, the infection-time distribution would be constructed based on the per-step incidence *S*_*k*_*h*_*k*_ rather than relying on the differences in *S*_*k*_. In this context, individual survival would be defined by the hazard *h*_*k*_ from the moment a person becomes susceptible. Furthermore, the final-size term would be replaced by a term representing the number of cases within the observation window, reflecting the person-time at risk. The only term that may remain unchanged would be the recovery-interval term. These extensions were not implemented here and are left for future work.

## 5 Limitations

The case studies highlight limitations of data fidelity and observational proxies. Not all event times used in the case studies were independently observed. In the H1N1 dataset, recovery times were generated using an exponential infectious period with a mean of 5.5 d, so that case study is best interpreted as a demonstration of the discrete inference framework under inherited assumptions, and the estimate of *γ* as a consistency check. It was retained as it provides a direct comparison with the survival-analytic approach of KhudaBukhsh et al. [12]. Additionally, in the Ebola analyses, hospitalisation was used as a proxy for removal. The inferred infectious period should therefore be read as a model-based removal interval tied to the observation process, and the *R*_0_ values are conditional on that definition and on the observation window. For the same reason, the population sizes *N* , *N*_*w*_ and *N*_0_–*N*_2_ are best read as effective at-risk sizes within the chosen model and window, not as total population sizes. The recording of a case is part of the same observation process. Cases missed at a constant rate are absorbed into these effective sizes (Section 2(c)(ii)), although in Hagelloch, where the class sizes were bounded, missed cases would also have lowered the estimated *R*_0_. However, if recording practices changed during an outbreak, for example as case finding improved, or if they depended on the duration or severity of a person’s illness, this would affect the infection-time curve or the recorded infectious periods. Such changes were not modelled here, and would be hard to separate from changes in transmission using the line list alone.

The transmission models rely on simplifying assumptions that DDSA shares with continuous-time DSA. Mean-field forces of infection average over individual-level heterogeneity in contacts, which can be a poor match when transmission is concentrated in households or classrooms. In Hagelloch, for example, the group-level model underpredicted the attack rates of Classes 0 and 2 by about 15% and brought the class epidemics closer together in time than observed. The analyses also assume a closed population over the observation window, which suits single outbreaks but not endemic transmission. The same assumptions affect how uncertainty is represented. The predictive bands reflect only parameter uncertainty, so they do not include day-to-day variation, which a chain-binomial version of the GLCT or Boxcar map would add. In addition, the infection-time likelihood treats infections as independent draws from the mean-field curve. In simulated epidemics, intervals for the infectious period, informed by individual recovery intervals, were generally close to nominal. Credible intervals for *R*_0_ were narrower than nominal even with the correct model. Nominal 95% intervals covered the true *R*_0_ in 17–80% of outbreaks, least often near *R*_0_ = 1, whereas coverage was close to nominal on data drawn independently from the mean-field curve (Supplementary Section S4). The *R*_0_ intervals of the Ebola waves, whose *R*_0_ is close to 1, may therefore understate the uncertainty. Methods that may correct the *R*_0_ intervals are outside the scope of this study.

The discrete-time formulation also carries a predictable step-size dependence. At Δ*t* = 1 d the DPH means exceed the corresponding continuous-time Erlang means, so estimates must be converted before they are compared with continuous-time ones. In simulations, the daily step also affected estimates in two ways (Supplementary Section S4.2). When continuous-time infectious periods were recorded by day, same-day removals became one-day intervals, which biased the mean infectious period upward by about 2%. When the epidemic grew quickly, *R*_0_ was biased upward. At *R*_0_ = 4, DDSA estimates exceeded those of continuous-time DSA by 7–9% by the end of the outbreak. A finer time step removed the first bias and reduced the second.

The GLCT map, built stage by stage, allows at most one stage transition per day, so *r* cannot exceed the shortest recorded interval, and a single short interval can set this limit. In simulations, this cap overstated the variability of the infectious period and biased *R*_0_ upward (Supplementary Section S4.3). The Boxcar map accepts a law without this minimum and so has no such cap, although its *R*_0_ intervals remained narrower than nominal at the daily step. Coxian or branched DPHs (Section 2(b)(ii)), in which an early exit or a branch with few stages can take the short intervals, may relax the cap and allow a less variable infectious period, but fitting them was left for future work.

Each analysis uses a single Δ*t*, so data recorded at different reporting intervals would need to be aggregated or entered as interval-censored terms. As the present case studies use uniform daily cadences, neither adjustment was necessary.

Finally, on the computational side, all DDSA analyses in the main text used one set of sampler settings (Section 2(d)), which met the convergence targets here but has not been tested more widely. Summing integer-valued parameters out of the likelihood is exact, but its cost grows with the summed support whenever the parameter changes the recurrence map. For instance, the Hagelloch model evaluates the map for each of the 40 combinations of *r* and *N*_2_ at every gradient, which makes it the most computationally demanding analysis. Other automatic-differentiation backends may perform differently, but these were not tested.

## 6 Conclusions

We have presented Discrete Dynamic Survival Analysis (DDSA), a discrete-time reformulation of dynamic survival analysis for calendar-day epidemic line lists. The DDSA likelihood has the same three components as continuous-time DSA (infection times, recovery intervals and final size), but each is a discrete analogue computed from a difference-equation map that replaces the continuous-time ODE. Infection times enter through a Multinomial distribution over calendar days, recovery intervals through discrete phase-type distributions, and the final size through a Binomial term. These components converge to their continuous-time counterparts as Δ*t* → 0, and each is computed exactly by iterating the map, without solver-based interpolation or custom numerical schemes for non-geometric infectious periods. They are differentiable in the continuous parameters, so gradient-based inference runs in standard probabilistic programming frameworks without model-specific numerical code.

Applied to three historical outbreaks of influenza, Ebola, and measles, under both homogeneous and group-structured compartmental models, DDSA accommodated incomplete line lists through composable likelihood terms for right-censored recoveries and missing onset times. Where published continuous-time DSA analyses were available (influenza and Ebola), its estimates were similar, except for the effective population size and reproduction number for H1N1. This agreement partly reflects shared modelling choices. Synthetic experiments showed that each type of data plays a different inferential role. Infection times determine the growth rate, the number never infected bounds *R*_0_ when the population at risk is known, and recovery intervals identify the infectious period. As a result, a misspecified infectious-period shape biased the estimates while leaving the incidence fit visibly unchanged. Only the recovery intervals revealed it. When recovery intervals were observed, the misspecification inflated the estimated reproduction number. Without them, it inflated the transmission rate and shortened the infectious period.

By keeping discrete-time inference aligned with the calendar-day structure of line-list data while remaining expressible in standard probabilistic programming tools, DDSA offers a transparent and extensible route to fitting mean-field epidemic models of single outbreaks to individual-level records.

## Data Availability

All data used in this study are openly available. Code and notebooks reproducing all analyses are archived on Zenodo (https://doi.org/10.5281/zenodo.23924832) and in the Julia package DiscreteDynamicSurvivalAnalysis.jl (https://github.com/epirecipes/DiscreteDynamicSurvivalAnalysis.jl).

https://doi.org/10.5281/zenodo.23924832

