## Supplementary material for "Discrete dynamic survival analysis of epidemic case line lists"

Supplementary Material for
### Discrete dynamic survival analysis of epidemic case line lists

Sandra Montes-Olivas, Sam Abbott, Sebastian Funk,
Michael B. Gravenor, Simon D.W. Frost

#### S1 Prior specifications

Tables S1 to S5 present the prior distributions and parameter bounds used in the main manuscript analyses. These tables are formatted as they appear in the `Turing.jl` `@model` blocks found in the accompanying notebooks (refer to Section S5). The integer-valued parameters ($N$, $r$) were summed out of the likelihood over their specified ranges, and their posterior draws were obtained from their conditional distributions after sampling. Unless otherwise noted, per-step parameters ($\gamma$, $p_\gamma$) refer to the one-day step of the discrete-time map.

##### S1.1 Influenza A(H1N1), Washington State University, 2009

This analysis implemented a single homogeneous-mixing SIR model with a geometric infectious period, as described in Section 3(c) of the main manuscript. The four parameters ($N$, $\beta$, $\gamma$ and $\rho$) were inferred jointly using practical-identifiability priors. $N$ was summed out of the Binomial final-size term.

Table S1: Priors for the H1N1 WSU case study. $K = 2{,}276$ is the observed number of cases.

| Parameter | Prior | Rationale / bounds |
|---|---|---|
| $N$ | DiscreteUniform($K$+1, 18,000) | At-risk pool: at least $K$+1; upper bound ≈ WSU student body (Vaidya et al. 2015). Summed out of the final-size term |
| $\beta$ | Uniform(0.01, 5.0) | Per-day transmission rate |
| $\gamma$ | LogNormal(log(1/5.5), 0.3) truncated to (0.01, 1.0) | $1/\gamma$ centred on the literature mean IP of 5.5 d (95% prior mass 3.0–9.9 d) |
| $\rho$ | Beta(1.0, 10.0) | Initial infected proportion (small seed) |

##### S1.2 Ebola virus disease, Democratic Republic of the Congo, 2018

The analysis applied the same SIR model, with a geometric infectious period, independently to each of the three waves (W1, W2, and W3), as described in Section 3(d) of the main manuscript. For each wave, the population at risk $N$ was characterised by a prior distribution defined as DiscreteUniform($K_{\text{total}}, K_{\text{total}} + 19,999$). As for H1N1, $N$ was summed out of the Binomial final-size term. In Waves 2 and 3, which started during ongoing transmission, the posteriors for $\rho$ were found to concentrate in the upper tail of its prior, indicating approximately 22 and

37 initial infectives, respectively. Across all three waves, these estimates were consistent with findings reported by Vossler et al. (2022).

Table S2: Priors for each Ebola DRC wave. $K_{\text{total}}$ denotes the total number of recorded cases in the wave and sets the lower bound of $N$.

| **Parameter** | **Prior** | **Rationale / bounds** |
|---|---|---|
| $\beta$ | Uniform(0.01, 1.0) | Per-day transmission rate |
| $\gamma$ | Uniform(0.01, 1.0) | Per-day recovery rate |
| $\rho$ | Beta(1.0, 500.0) | Initial infected fraction |
| $N$ | DiscreteUniform($K_{\text{total}}$, $K_{\text{total}}$+19,999) | Summed out of the final-size term. Lower bound = observed cases in wave; upper bound above the shared $N \approx 16{,}000$ reported by Vossler et al. (2022) |

##### S1.3 Measles, Hagelloch, Germany, 1861

This analysis implemented a three-group stratified SIR model with four transmission parameters and a Negative Binomial DPH infectious period, as described in Section 3(e) of the main manuscript. The GLCT and Boxcar variants shared the same priors. The upper bound of each class size, $(N_{0,\text{max}}, N_{1,\text{max}}, N_{2,\text{max}}) = (99, 39, 77)$, was the 197 children under 14 recorded by Pfeilsticker in 1861 (as cited by Neal and Roberts 2004) minus the observed cases in the other two classes, $(K_0, K_1, K_2) = (90, 30, 68)$. These bounds did not jointly restrict $N_0 + N_1 + N_2$ to 197. The class sizes and the number of stages $r$ were summed out of the likelihood. $N_2$, which also set the seed fraction $\rho_2 = 1/N_2$, was summed jointly with $r$ over their 40 combinations ($r = 1$–4, $N_2 = 68$–77). The upper bound $r_{\text{max}} = 4$ was the shortest recorded infectious period, so this interval set the range of shapes the model could fit (Section S4).

##### S1.4 Data-availability synthetic scenarios

These analyses used a synthetic line list generated by a GLCT simulator with four geometric stages (a discretised Erlang-4 infectious period), with $\beta = 0.5\,\text{day}^{-1}$, $N = 99$, $\rho = 0.01$ and $T = 40$ days, as described in Section 3(a) of the main manuscript. Each of the $r = 4$ stages discretised an exponential stage with rate $\lambda = 1\,\text{d}^{-1}$ (continuous-time mean 4 d) at $\Delta t = 1\,\text{d}$, so the per-step progression probability was $g = 1 - e^{-\lambda \Delta t} \approx 0.632$ and the DPH mean was $\overline{W} = r/g \approx 6.33\,\text{d}$. This mean exceeded 4 d because each geometric stage had an expected stay of $1/g \approx 1.58\,\text{d}$ rather than $1/\lambda = 1\,\text{d}$. As $\Delta t \to 0$, $g \approx \lambda \Delta t$ and $\overline{W} \to r/\lambda$. The data were generated by this discrete map, so 6.33 d was the true mean infectious period of the line list. The deterministic map gave the attack rate $\tau$ and the infection-time distribution $\mathbf{f}$. The line list was then drawn as $K \sim \text{Binomial}(N, \tau)$, daily infection counts $\mathbf{y} \sim \text{Multinomial}(K, \mathbf{f})$ and recovery intervals $\delta_i$ from the DPH probability mass function. The NegBin DPH (GLCT), geometric, and Boxcar models used the same prior families across data-availability scenarios A (full line list), B (infection times only), and C (recovery intervals only). In Scenario C, only the NegBin DPH and geometric models were fitted. Without infection times, only the recovery-interval term remained, and the NegBin likelihood was the same for the GLCT and Boxcar maps. In Scenario B, the NegBin DPH and Boxcar models sampled $R_0$ and the mean infectious period $\mu$, with $p_\gamma = r/\mu$ and $\beta = R_0/\mu$, so that these parameters kept their meaning across values of $r$.

Table S3: Priors for the Hagelloch measles case study (shared between the GLCT and Boxcar variants). $\delta_{\text{floor}}$ is the minimum observed infectious-period duration $\delta$ (4 days), the start of the support of the offset NegBin DPH ($\delta_{\text{floor}} - r$ deterministic stages followed by $r$ geometric stages).

| **Parameter** | **Prior** | **Rationale / bounds** |
|---|---|---|
| $N_0$ | DiscreteUniform(90, 99) | Pre-school class size; lower bound = observed $K_0$, upper bound from census; summed out |
| $N_1$ | DiscreteUniform(30, 39) | Younger-school class size; bounds as above; summed out |
| $N_2$ | DiscreteUniform(68, 77) | Older-school class size; bounds as above; summed out jointly with $r$ |
| $\beta_{C_1}$ | Exponential(0.2) | Within-class transmission rate, Class 1 (younger school) |
| $\beta_{C_2}$ | Exponential(0.1) | Within-class transmission rate, Class 2 (older school) |
| $\beta_H$ | Exponential(0.1) | School-to-pre-school transmission rate (household proxy) |
| $\beta_G$ | Exponential(0.1) | Global transmission rate, acting on all groups |
| $\overline{\delta}$ | Uniform($\delta_{\text{floor}} + 0.5$, 25.0) | Mean infectious period in days |
| $r$ (NegBin phases) | DiscreteUniform(1, $r_{\max}$) | Phase count; $r_{\max} = \min(\delta) = 4$; summed out jointly with $N_2$ |
| $p_\gamma$ | deterministic $= r/(\overline{\delta} - \delta_{\text{floor}} + r)$ | Implied per-stage transition probability |

#### S1.5 DDSA versus count-based inference

This comparison used a synthetic line list from the same simulator as in Section S1.4 ($\beta = 0.5\,\text{day}^{-1}$, $N = 99$, $\rho = 1/N$, and $T = 40\,\text{days}$), as described in Section 3(b) of the main manuscript. The DDSA model was fitted to the full line list. The four count-based comparators were SIR ODE models fitted to the daily incidence only, with continuous-time rates $\beta$ and $\gamma$ and initial infected fraction $i_0$. The Negative Binomial observation model had a mean equal to the ODE incidence and dispersion $\phi$.

Table S4: Priors for the data-availability synthetic study. "Scen" indicates the scenarios in which each prior applies (A: full data; B: infection-only; C: recovery-only). $r_{\text{max}}$ is the prior upper bound on the discrete phase count: $r_{\text{max}} = 10$ in Scenarios A and B and $r_{\text{max}} = \min(\delta) = 4$ in Scenario C. In Scenario A, values of $r$ above $\min(\delta)$ have zero likelihood, because a NegBin DPH with $r$ phases cannot produce intervals shorter than $r$ days. In Scenario B, $p_\gamma = r/\mu < 1$ was required, so the effective joint prior of $(r, \mu)$ was the product of the two uniforms restricted to $\mu > r$.

| **Parameter** | **Prior** | **Model(s)** | **Scen** |
|---|---|---|---|
| $\beta$ | Uniform($10^{-4}$, 2.5) | NegBin DPH, Boxcar, Geometric | A |
| $\beta$ | Uniform($10^{-4}$, 2.5) | Geometric | B |
| $R_0$ | Uniform(0.5, 5.0) | NegBin DPH, Boxcar | B |
| $\rho$ | Beta(1.0, $N-1$) | All models | A, B |
| $p_\gamma$ (= `nb_p`) | Beta(1.0, 1.0) | All models fitted (A, C); Geometric (B) | A, B, C |
| $\mu$ (mean IP, days) | Uniform(1.0, 30.0) | NegBin DPH, Boxcar | B |
| $r$ (= `nb_r`) | DiscreteUniform(1, $r_{\text{max}}$), summed out | NegBin DPH (A, B, C), Boxcar (A, B) | A, B, C |
| $r$ | $\equiv 1$ | Geometric (misspecified) | A, B, C |

Table S5: Priors for the comparison of DDSA with count-based inference. The four count models scored the daily incidence with a Poisson or Negative Binomial (NB) observation model. They combined an exponential infectious period with $\gamma$ free and Poisson counts, an exponential infectious period with $\gamma$ free and NB counts, an Erlang-4 infectious period with $\gamma$ free and NB counts, and an exponential infectious period with $\gamma$ fixed at $1/4\,\text{day}^{-1}$ and NB counts.

| **Parameter** | **Prior** | **Model(s)** |
|---|---|---|
| $\beta$ | Uniform($10^{-4}$, 2.5) | All five models |
| $\rho$ | Beta(1.0, $N-1$) | DDSA |
| $p_\gamma$ (= `nb_p`) | Beta(1.0, 1.0) | DDSA |
| $r$ (= `nb_r`) | DiscreteUniform(1, $\min(10, \min\delta)$), summed out | DDSA |
| $i_0$ | Uniform($10^{-4}$, 0.2) | Count models |
| $\gamma$ | Uniform(0.02, 1.0) | Count models with $\gamma$ free |
| $\phi$ | Exponential(5.0). NB variance $m + m^2/\phi$, where $m$ is the expected daily incidence | Count models with NB counts |

#### S2 Identifiability and information in the data

In Section 3(b) of the main manuscript, the count-based models, fitted only to the daily incidence counts, left $R_0$ almost as uncertain as its prior when the removal rate was estimated, whereas DDSA, fitted to the full line list, did not. Several factors differed between the two approaches: the forward model (discrete map or continuous-time ODE), the observation likelihood, the use of recovery intervals, and the priors. In this section, one factor was changed at a time to locate the step at which $R_0$ stopped being identified, and the likelihood term responsible. Results that this supplement summarises without a table or figure are shown in the repository's supplementary guide notebook (Section S5) and are cited by its section and heading, for example (guide, S2: *The attack-rate sweep*).

$N$ denotes the whole population at risk, including the initial infectives, as in the main manuscript. Where the initially susceptible and initially infectious individuals are specified separately, or a continuous-time DSA likelihood is written in its original notation (KhudaBukhsh et al. 2020), $n$ is the number of initially susceptible individuals and $m$ the number of initial infectives, so $N = n + m$.

**Components of DDSA.** DDSA consists of a likelihood, a discrete-time forward map and one of two infectious-period structures. Each result in Sections S2–S4 is attributed to one of these, or to the model assumptions that DDSA shares with continuous-time DSA:

- (L) The *DDSA likelihood*: Multinomial infection-time term, Binomial final-size term and recovery-interval term. It has the same structure as the continuous-time DSA likelihood, with infection times treated as independent draws from the mean-field curve, evaluated on daily steps.
- (M) The *discrete-time forward map* on steps of $\Delta t$ (one day in the main manuscript), common to both infectious-period structures.
- (G) The *GLCT map*: a NegBin DPH infectious period built from $r$ geometric stages, with at most one stage transition per step (the serial, stage-by-stage discretisation used throughout).
- (B) The *Boxcar map*: an infection-age map that accepts any discrete interval law.

With a geometric infectious period ($r = 1$), the GLCT and Boxcar maps coincide. DDSA and continuous-time DSA both assume a closed, homogeneous, mean-field population.

##### S2.1 Design

All fits used the synthetic outbreak of Sections 3(a) and 3(b) of the main manuscript (Section S1.4), with $N = 99$ including one initial infective (98 initially susceptible). The outbreak had $K = 91$ cases (attack rate 0.92), and the true values were $\overline{W} = 6.33\,\text{d}$ and $R_0 = 3.164$. These data were drawn from the DDSA working model, so cases were independent given the parameters. Sections S3 and S4 used stochastic epidemics. Every model used the same priors, $R_0 \sim \text{U}(0.5, 5)$, mean infectious period $\mu \sim \text{U}(1, 30)\,\text{d}$, $\rho \sim \text{Beta}(1, N-1)$ and $r \sim \text{U}\{1, \ldots, 10\}$ (summed out, $p = r/\mu$), with $N = 99$ fixed, and NUTS with four chains of 4,000 draws started from prior draws.

**The six models.** *Infection-only DDSA* corresponds to Scenario B of Section 3(a) of the main manuscript: the discrete map, with the Multinomial infection-time term and the Binomial final-size term. *Map–Poisson* and *map–NegBin* kept the discrete map and scored the daily incidence counts with independent Poisson or Negative Binomial terms. *ODE–Poisson* and *ODE–NegBin* also replaced the discrete map with an Erlang-$r$ ODE, the continuous-time analogue of the

GLCT structure (G) used by the other models. These four are the count models. *Full-data DDSA* corresponds to Scenario A: infection-only DDSA plus the recovery-interval term $\ell_R$, which requires $r \le \min_i \delta_i$ (Section S4.3; Table S6).

Table S6: The identifiability ladder: posterior medians [95% CrI] of $R_0$ and the mean infectious period (IP) for the synthetic outbreak of Sections 3(a)–(b), with $N = 99$ known and the six models defined in the text. Panel (a): common priors. Every fit met $\hat{R} \le 1.01$ with no divergent transitions. Panel (b): wider prior on $R_0$, with the largest $\hat{R}$, the smallest bulk ESS and the number of divergent transitions over the four chains. $^\dagger$: did not meet these targets. Truth: $R_0 = 3.16$, mean IP 6.33 d (DPH mean). For the ODE models, the mean IP is the continuous Erlang mean.

*(a) Common priors,* $R_0 \sim \mathrm{U}(0.5, 5)$

| **Model** | **Observation likelihood** | $\ell_R$ | $R_0$ | **Mean IP (d)** |
|---|---|---|---|---|
| Infection-only DDSA | Multinomial + Binomial (Scenario B) | – | 3.11 [2.44, 4.44] | 6.34 [3.76, 11.2] |
| Map–Poisson | Poisson daily counts | – | 4.15 [2.42, 4.97] | 8.84 [4.05, 12.2] |
| Map–NegBin | Negative Binomial daily counts | – | 4.19 [2.43, 4.97] | 8.77 [4.08, 12.2] |
| ODE–Poisson | Poisson daily counts | – | 4.24 [2.56, 4.97] | 10.1 [5.02, 13.5] |
| ODE–NegBin | Negative Binomial daily counts | – | 4.25 [2.54, 4.97] | 9.93 [4.83, 13.6] |
| Full-data DDSA | Multinomial + Binomial (Scenario A) | yes | 3.13 [2.74, 3.57] | 6.40 [6.03, 6.83] |

*(b) Wider prior,* $R_0 \sim \mathrm{U}(0.5, 20)$

| **Model** | $R_0$ | **Mean IP (d)** | $\hat{R}$ | **Bulk ESS** | **Divergent** |
|---|---|---|---|---|---|
| Infection-only DDSA | 3.20 [2.46, 10.3] | 6.70 [3.77, 27.7] | 1.005 | 757 | 0 |
| Map–Poisson$^\dagger$ | 10.1 [3.65, 14.6] | 22.5 [7.46, 29.7] | 1.009 | 536 | 10 |
| Map–NegBin$^\dagger$ | 10.2 [3.93, 15.1] | 22.4 [7.67, 29.6] | 1.007 | 513 | 25 |
| ODE–Poisson$^\dagger$ | 9.16 [3.11, 12.9] | 23.0 [6.83, 29.6] | 1.03 | 169 | 65 |
| ODE–NegBin | 9.64 [3.96, 13.5] | 23.7 [8.93, 29.7] | 1.003 | 875 | 0 |
| Full-data DDSA | 3.13 [2.75, 3.58] | 6.40 [6.04, 6.83] | 1.001 | 7,695 | 0 |

#### S2.2 Information from the never-infected count

In this high-attack-rate outbreak, replacing the Binomial final-size term with independent Poisson counts substantially weakened inference on $R_0$. Changing the forward model or adding overdispersion to the counts did not restore it (Table S6a). Independent Poisson daily counts factorise exactly into the DDSA timing term and a Poisson final-size term,

$$\sum_t \log \mathrm{Pois}(y_t; \mu_t) = \log \mathrm{Multinomial}(\mathbf{y}; K, \mathbf{f}) + \log \mathrm{Pois}(K; N\tau), \qquad \mathbf{f} = \boldsymbol{\mu} / \textstyle\sum_t \mu_t, \quad (1)$$

so map–Poisson and infection-only DDSA, evaluated on the same map, use the infection days in the same way and differ only in how the number of cases is scored, $\mathrm{Pois}(K; N\tau)$ against $\mathrm{Binomial}(K; N, \tau)$. For the Negative Binomial and ODE count models, the comparison in Table S6 is empirical. The Poisson factor loses the observation that the $N - K$ individuals at risk were not recorded as incident cases during the observation window (the never-infected cell), which the Binomial term carries through $(N - K)\log(1 - \tau)$. Under the DDSA convention, this cell also contains the initial infectives, $1 - \tau = \rho + S_T$. The Fisher information on $\tau$, a measure of how precisely the number of cases determines $\tau$, is $N/\{\tau(1 - \tau)\}$ under the Binomial term and $N/\tau$ under the Poisson term. In large samples, the posterior SD of $\tau$ is roughly the inverse square root of this information, so the Poisson term should give an SD larger by a factor of $\sqrt{1/(1 - \tau)}$, about 1.6 at an attack rate of 0.6 and 2.2 at 0.8. Near $\tau = 1$, this approximation no longer holds. In this example, the Binomial term excluded complete infection, whereas the Poisson factor changed little as $\tau$ approached one, letting the posterior extend toward larger $R_0$. Table S7 isolates this final-size factor (guide, S2: *Table S7: isolating the final-size term*).

Table S7: Isolating the final-size term: posterior medians [95% CrI] of $R_0$. All fits used infection-only DDSA data (infection times and final size) and $R_0 \sim \mathrm{U}(0.5, 5)$; $N = 99$ is known unless a prior on $N$ is given. True $R_0 = 3.16$ (first row). $^{\ddagger}$: numerical grid posterior (Section S5). The other fits used NUTS.

| **Likelihood** | $R_0$ |
|---|---|
| *Truth* | 3.16 |
| Multinomial timing + Binomial final size (= infection-only DDSA) | 3.11 [2.44, 4.44] |
| Poisson daily counts (= map–Poisson) | 4.15 [2.42, 4.97] |
| Counts with a never-infected cell, Multinomial($N; \tau\mathbf{f}, 1-\tau$), discrete map | 3.12 [2.43, 4.44] |
| Counts with a never-infected cell, continuous-time ODE | 3.14 [2.46, 4.46] |
| Infection-only DDSA with $N \sim \mathrm{U}(91, 500)$ summed out | 4.20 [2.31, 4.96] |
| Infection-only DDSA with $N \sim \mathrm{U}(91, 20{,}090)$, $\rho \sim \mathrm{Beta}(1, 98)^{\ddagger}$ | 4.18 [2.23, 4.98] |
| Infection-only DDSA with $N \sim \mathrm{U}(91, 20{,}090)$, seed count $I_0 = \rho N \sim \mathrm{Exp}(1)^{\ddagger}$ | 4.18 [2.23, 4.98] |

**A never-infected cell restores the information.** A count model with one extra cell for the never-infected, Multinomial($N; \tau f_1, \ldots, \tau f_T, 1-\tau$) on $(h_1, \ldots, h_T, N-K)$, is algebraically the DDSA infection-time plus final-size likelihood,

$$\mathrm{Multinomial}\big((\mathbf{h}, N-K); N, (\tau\mathbf{f}, 1-\tau)\big) = \mathrm{Binomial}(K; N, \tau)\,\mathrm{Multinomial}(\mathbf{h}; K, \mathbf{f}). \quad (2)$$

Fitted to aggregated daily counts, it gave similar posterior summaries under the discrete map and the continuous-time ODE (Table S7). In this setting, without recovery intervals, much of the information on $R_0$ came from the count of individuals at risk who were never infected. This information did not require individual records or a discrete-time forward model. This count model is equivalent to the count-data DSA likelihood of Roy et al. (2026), $\ell = s_T^{N-K} \prod_j (s_{j-1} - s_j)^{Y_j}$, up to a parameter-free multinomial coefficient, when its cell probabilities are expressed per initially susceptible individual. DDSA instead expresses them per member of the whole population $N$.

#### S2.3 Prior sensitivity and the growth-rate ridge

Without recovery intervals, the count-model posteriors remained sensitive to the prior bounds: widening the $R_0$ prior shifted them along the $R_0$–mean-IP ridge until the upper bound on the mean infectious period became limiting (Table S6; Figure S1). Infection-only DDSA constrained $R_0$ more strongly, but its upper tail remained sensitive to the prior in this small outbreak. This tail was not seen in the larger populations (Table S8). The broad count-model intervals of Section 3(b) of the main manuscript reflected the same weak identification, widened by the long upper tail of the $R_0$ prior implied by uniform priors on $(\beta, \gamma)$.

**Infection timing constrains the growth rate.** The early growth rate of the incidence depends on both $R_0$ and the infectious period, so a larger $R_0$ with a longer infectious period can give the same growth. The growth rate implied by the posterior draws was close to the truth for the three models of Figure S1, but $R_0$ and the mean infectious period traded off along this ridge. In this outbreak, infection-only DDSA spread along the ridge almost as widely as map–Poisson, but its posterior was centred closer to the truth (Table S6). Recovery intervals

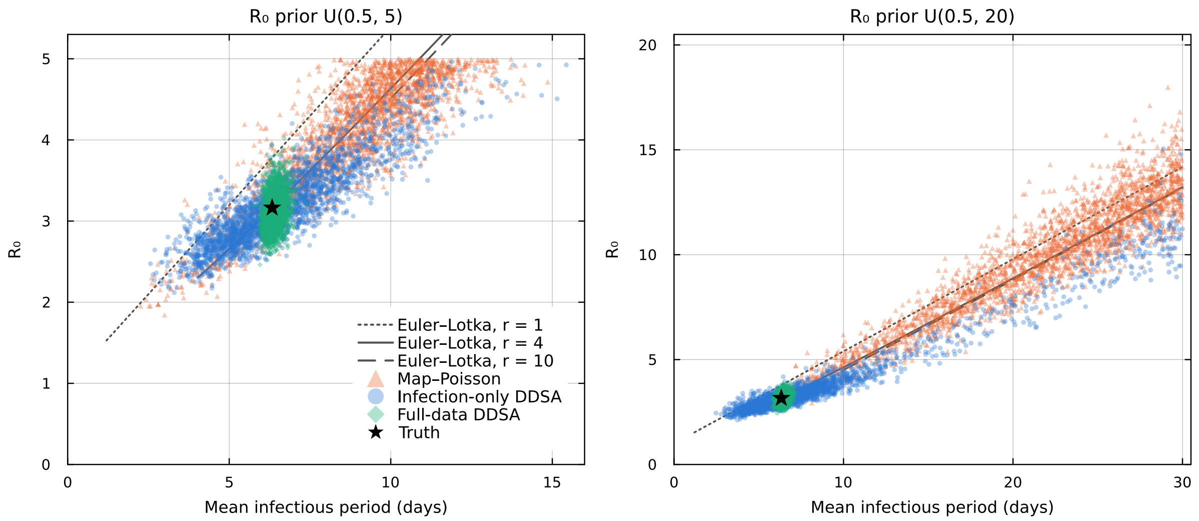


Figure S1: Joint posterior draws of $R_0$ and the mean infectious period for infection-only DDSA (Scenario B), map–Poisson and full-data DDSA (Scenario A), under $R_0 \sim \mathrm{U}(0.5, 5)$ (left) and $\mathrm{U}(0.5, 20)$ (right). Grey lines: pairs of $R_0$ and mean infectious period that give the true early growth rate (discrete Euler–Lotka relation; Diekmann et al. 2021), for $r = 1, 4, 10$. Star: truth.

constrained the posterior position along the ridge much more strongly. Draw-wise growth rates and correlations are in the repository's supplementary guide (guide, S2: *Figure S1: the growth-rate ridge*).

**The infectious-period shape.** Without recovery intervals, the shape of the infectious period was weakly informed: the posterior of $r$ moved little from its effective prior (uniform, restricted to $\mu > r$). The posterior probabilities of each value of $r$ are in the same section of the repository's supplementary guide. Full-data DDSA concentrated on $r = 4$, but this also reflected a restricted support. The data gave $r < 4$ negligible posterior weight, and the serial DPH of the GLCT map (G) gave zero likelihood to $r$ above the shortest recorded interval, which here equalled the true $r$ ($\min_i \delta_i = 4$). This concentrated posterior should therefore not be read as unrestricted identification of the shape. With short or rounded intervals, the cap can bind below the truth (Section S4).

#### S2.4 Attack rate, population size and an unknown $N$

The advantage of the Binomial final-size term depended on the attack rate and the population size. Infection-only DDSA and map–Poisson were refitted to outbreaks with $R_0 \approx 1.5$, 2 and 3.16 at $N = 99$ and 1,000, as numerical grid posteriors (guide, S2: *The attack-rate sweep*). In the lower-attack-rate settings examined, map–Poisson also constrained $R_0$, although less precisely. Where its posterior of $\tau$ stayed well below 1, the posterior SD of $\tau$ was larger than under infection-only DDSA by close to the factor $\sqrt{1/(1-\tau)}$ of Section S2.2. The difference grew sharply once the Poisson posterior could reach $\tau \to 1$, because $R_0$ rises steeply as $\tau$ approaches 1. The loss of information in the main example should therefore not be generalised to all incidence-only analyses.

**An unknown population size.** With infection-only data and $N$ unknown, infection-only DDSA gave posterior summaries similar to map–Poisson (Table S7), because summing over $N$ removes most of the information in the never-infected cell. With $\tau$ fixed and constant weights over $N \geq K$, the summed final-size term is $\sum_{N \geq K} \binom{N}{K} \tau^K (1-\tau)^{N-K} = 1/\tau$, which, like the Poisson factor, changes little as $\tau$ approaches 1. The fitted priors were bounded, so this identity illustrates the mechanism only, and the fitted results depended on the priors. With a wide support and the seed prior on the fraction scale, two of the four NUTS chains stayed in a subcritical local mode, with about 91 initial infectives and a long infectious period. A numerical grid posterior gave this mode negligible mass and the same $R_0$ as the seed prior on the count scale (Table S7). Section S3 compares summing $N$ out with the imputation step of continuous-time DSA on the WSU line list, which has recovery intervals.

**Numerical grid posteriors.** After $r$ (and $N$) were summed out, the fits without recovery intervals had three real-valued parameters ($R_0$, $\mu$, $\rho$). Where NUTS mixed poorly, their posteriors were evaluated on grids with a midpoint rule. This was the case for the infection-only fits of the attack-rate sweep at $N = 1{,}000$, whose mean infectious period was bimodal, and for the larger-population fits without recovery intervals at every $N$, because NUTS did not reach $\hat{R} \leq 1.01$ at $N \geq 1{,}000$. The grid checks are listed in Section S5. The full-line-list fits showed no such multimodality and were sampled with NUTS.

#### S2.5 Larger populations

The ladder used one outbreak of 99 people, almost all of whom were infected. To see how its conclusions changed with population size, four models were fitted to one outbreak at each of $N =$ 99, 1,000 and 10,000, with the same $R_0$ and infectious period, and their fitted trajectories were compared with the true mean-field trajectory (Table S8, Figure S2). Map–Poisson, infection-only DDSA and full-data DDSA added one source of information at a time, first the Binomial final-size term (Eq. (1)) and then the recovery intervals. The fourth was ODE–Poisson.

All three datasets were drawn from the DDSA working model, like the ladder outbreak (the $N = 99$ dataset), with a recovery interval from the four-stage DPH for each case and the initial infective, so every interval was at least 4 d. Because the cases were independent given the parameters, the comparison concerned what the data identify. The priors were those of the ladder, with $R_0 \sim \mathrm{U}(0.5, 20)$ and $\rho \sim \mathrm{Beta}(1, N-1)$. The ODE summed $r$ over $1, \ldots, 40$ without the constraint $\mu > r$, which only the discrete map needs. Full-data DDSA was sampled with NUTS as in the ladder ($\hat{R} \leq 1.001$, no divergent transitions), and the other three models were evaluated as numerical grid posteriors. With a single dataset at each $N$, the results illustrate how the models behave.

**What each source of information adds.** Scoring the never-infected cell improved inference on $R_0$, and the recovery intervals further reduced the uncertainty in the parameters and trajectories (Table S8, Figure S2). Infection-only DDSA placed the median of $R_0$ close to the truth at each $N$ and was more precise than map–Poisson in the largest outbreak. Both count models moved towards the true $R_0$ as $N$ grew, but their infectious-period estimates differed. The ODE was misspecified for these data from the discrete map and fitted a longer infectious period. Its rows therefore show how the count models behave under this generator, not which time scale is more accurate.

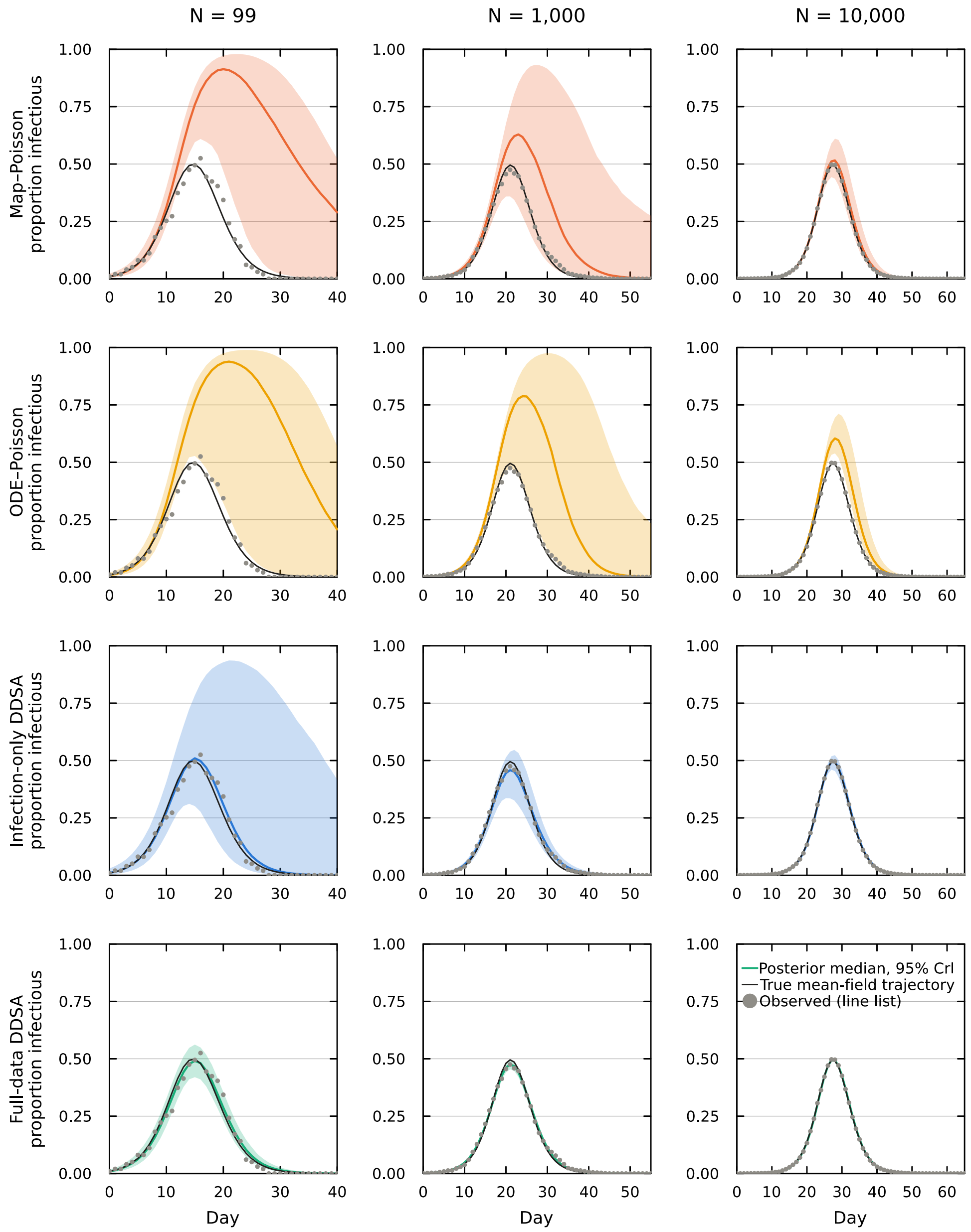


Figure S2: Proportion infectious, $I(t)/N$, for one simulated outbreak at each population size (columns: $N$ = 99, 1,000 and 10,000) under the four models of Table S8 (rows). Coloured line and band: posterior median and 95% CrI of the expected proportion infectious (parameter uncertainty only). Black line: true mean-field trajectory. Grey points: proportion infectious counted from the line list. Expected new infections per day are in the repository's supplementary guide (guide, S2: *Table S8 and Figure S2: larger populations*).

Table S8: Larger populations: posterior medians [95% CrI] of $R_0$ and the mean infectious period, and the peak of the expected number infectious, for one outbreak at each population size. Full-data DDSA was sampled with NUTS and the other models were evaluated as numerical grid posteriors. For ODE–Poisson, the mean infectious period is the continuous-time mean (within 0.1 d of its mean recorded by day), and the posterior of $r$ was still rising at the upper end of its support ($r = 40$).

| $N$ | Model | $R_0$ | Mean infectious period (d) | Peak number infectious |
|---|---|---|---|---|
| 99 | Truth | 3.16 | 6.33 | 49 |
| | Map–Poisson | 10.16 [3.93, 14.66] | 22.62 [8.26, 29.62] | 91 [61, 97] |
| | ODE–Poisson | 8.84 [2.98, 12.70] | 22.38 [6.99, 29.62] | 93 [52, 98] |
| | Infection-only DDSA | 3.20 [2.46, 10.28] | 6.69 [3.86, 27.38] | 51 [31, 93] |
| | Full-data DDSA | 3.13 [2.75, 3.58] | 6.40 [6.04, 6.83] | 49 [42, 56] |
| 1,000 | Truth | 3.16 | 6.33 | 495 |
| | Map–Poisson | 4.60 [2.72, 12.46] | 10.69 [4.86, 28.88] | 631 [360, 932] |
| | ODE–Poisson | 5.12 [2.75, 10.60] | 14.04 [6.71, 29.38] | 789 [473, 976] |
| | Infection-only DDSA | 3.15 [2.93, 3.42] | 6.44 [4.91, 7.74] | 458 [336, 546] |
| | Full-data DDSA | 3.05 [2.93, 3.17] | 6.44 [6.32, 6.57] | 475 [454, 496] |
| 10,000 | Truth | 3.16 | 6.33 | 4,928 |
| | Map–Poisson | 3.29 [2.95, 3.87] | 6.66 [5.61, 8.24] | 5,156 [4,429, 6,104] |
| | ODE–Poisson | 3.27 [2.94, 3.93] | 8.04 [7.04, 9.91] | 6,041 [5,387, 7,113] |
| | Infection-only DDSA | 3.18 [3.10, 3.25] | 6.34 [5.91, 6.69] | 4,942 [4,599, 5,235] |
| | Full-data DDSA | 3.17 [3.13, 3.21] | 6.33 [6.29, 6.37] | 4,936 [4,865, 5,008] |

### S3 Validation against continuous-time DSA

Continuous-time DSA (KhudaBukhsh et al. 2020; Di Lauro et al. 2022) is the established method for fitting mean-field epidemic models to line lists. In this section, DDSA was validated against it in three steps. The re-implementations of continuous-time DSA were first compared with the published codes of KhudaBukhsh et al. (2020) and Di Lauro et al. (2022). DDSA was then compared with these re-implementations on simulated outbreaks with known parameters. Finally, the case-study estimates were compared with the published continuous-time analyses of the same data.

#### S3.1 Reference implementations

The continuous-time re-implementations were written in Julia from the equations and algorithms of the published papers: the SDS likelihood of KhudaBukhsh et al. (2020, eq. 3.3) with a fourth-order Runge–Kutta mean field, their Algorithm 4.1 for an unknown population size, and the mean-field renewal equation for a Gamma infectious period, which solves the age-structured PDE of Di Lauro et al. (2022) along its characteristics. The priors and sampler of DDSA, stated in Section S3.2, were implemented so the comparison differed mainly in the model timescale and data resolution. In the example scripts distributed with the SDS code of KhudaBukhsh et al. (2020), with the priors of those scripts, the re-implementations produced posterior summaries similar to those of that code (Table S9; guide, S3: *Table S9: published codes in their own settings*). For the published Python code, which fits by maximum likelihood, its likelihood of infection times was compared with that of the re-implementation at the maximum found by the re-implementation, and the two agreed closely (Table S9). This check did not cover its optimisation procedure or its other likelihood components.

Table S9: The continuous-time re-implementations against the published codes of KhudaBukhsh et al. (2020; SDS, R) and Di Lauro et al. (2022; DSA, Python), on the examples distributed with those codes. SDS rows: ranges over outbreaks of the posterior median $R_0$, or medians [95% CrI], from four chains with the Gamma(0.001, 0.001) priors of the examples, with $R_0 = \beta(1+\rho_{\text{SDS}})/\gamma$ on the whole-population scale of DDSA. In the two outbreaks outside the stated agreement, $N$ was weakly identified ($\hat{R}$ of the published code 1.07–1.08). DSA row: largest difference in the likelihood of infection times $\ell_I$ at the re-implementation's maximum, with their PDE on the 10,000-point grid of their study and their initial condition.

| Setting | Published code | Re-implementation | Agreement |
|---|---|---|---|
| SDS, Example 2: 10 outbreaks, $n = 1{,}000$ known | $R_0$ 1.28–1.54 | $R_0$ 1.28–1.54 | $R_0$ medians within 0.003 |
| SDS, Example 2: the same outbreaks, $n$ imputed (Algorithm 4.1) | $R_0$ 1.01–1.81 | $R_0$ 0.98–1.81 | $R_0$ within 0.02 and $N$ within 2% in 8 of 10 |
| SDS, example 1: WSU, $n$ imputed | $R_0$ 1.09 [1.06, 1.14], $N$ 7,652 | $R_0$ 1.09 [1.05, 1.14], $N$ 7,734 | – |
| DSA, synthetic study: 5 outbreaks, $N = 10{,}000$, infection times only | $\ell_I$ at the re-implementation's optimum | $\ell_I$ at the same optimum | within 0.05 log units |

#### S3.2 Agreement on simulated outbreaks

Outbreaks were simulated from a continuous-time stochastic SIR model by exact event-driven simulation, following the validation design of KhudaBukhsh et al. (2020, §3.4): $n$ susceptibles and $m = 0.05\,n$ initial infectives, infection hazard $\beta SI/(n + m)$, and exponential infectious periods with mean 5 d. The grid was $n \in \{100, 1{,}000, 10{,}000\} \times R_0 \in \{1.5, 2, 4\}$ with 50 replicates per cell, not conditioned on a major outbreak, and each outbreak was observed up to a cut-off near the peak or near the end of the mean-field epidemic.

DDSA with a geometric infectious period was fitted to daily records, with infection day $\lfloor t \rfloor + 1$ and interval equal to the recovery day minus the infection day (one day for a same-day recovery), right-censored at the cut-off. With $r = 1$, there was no number of stages to cap. Daily recording instead set a minimum interval of one day, which affected the estimated mean infectious period (Section S4.2). The re-implemented continuous-time DSA was fitted to the exact times with $n$ known. Both used $\beta \sim \mathrm{U}(10^{-4}, 2.5)\,\mathrm{d}^{-1}$, $\mu \sim \mathrm{U}(1, 30)$ d and $\rho \sim \mathrm{Beta}(1, 10)$, and NUTS with four chains of 1,000 draws after 1,000 warm-up. The complete-data posterior, given every infection and recovery time of the simulated outbreak, was used as the reference for what each outbreak could identify.

The study comprised 3,150 Bayesian fits, including the arm on daily records spread within the day reported in the repository's supplementary guide and the step-refinement arms of Section S4.2. All fits were kept, including 7 with $\hat{R} > 1.01$ and 106 with at least one divergent transition. Excluding the fits with either warning changed the reported $R_0$ bias by at most 3.3 percentage points and coverage by at most 0.03 (Tables S10 and S13; guide, S3: *Paired comparison and convergence*).

DDSA and the re-implemented continuous-time DSA were fitted to the same outbreaks, so their paired difference largely removed the variation between outbreaks that affects both. For $R_0 \le 2$ and $n \ge 1{,}000$, the two posterior medians differed by less than 1% of $R_0$ at both cut-offs, within about one posterior SD (Table S10). At $n = 100$, they differed by 2–3%, less than their departures from the truth. The complete-data reference shared these departures, which reflect how small outbreaks, including minor ones, vary around the mean field. At $R_0 = 4$, DDSA gave higher estimates than continuous-time DSA, by 7–9% at the end cut-off and by 1–5% at the peak. Refining the records and the map reduced this difference (Section S4.2). Outside these settings, the two methods had similar interval coverage (guide, S3: *Table S10: agreement on*

Table S10: $R_0$ estimated by DDSA and by continuous-time DSA on the simulated outbreaks of Section S3.2 ($n$ initially susceptible), observed to a cut-off near the end (panel a) or near the peak (panel b). Bias: relative bias of the posterior median (%), mean over replicates. Complete data: posterior given every infection and recovery time. DDSA − DSA (exact): difference between the two posterior medians for the same outbreak, as a percentage of the true $R_0$ (mean, with its 95% CI) and in posterior SDs of DSA (exact) (median). 50 replicates per row, all fits included. Interval coverage, the arm on daily records spread within the day and the mean infectious period are in the repository's supplementary guide (guide, S3: *Table S10: agreement on simulated outbreaks* and guide, S3: *Mean infectious period*).

| | | Bias of the posterior median (%) | | | DDSA − DSA (exact) | |
|---|---|---|---|---|---|---|
| $n$ | True $R_0$ | Complete data | DDSA | DSA (exact) | % of $R_0$ [95% CI] | Posterior SDs |
| *(a) Cut-off near the end* | | | | | | |
| 100 | 1.5 | −8.8 | −10.2 | −12.8 | +2.6 [1.5, 3.6] | 0.2 |
| 100 | 2 | −11.3 | −10.5 | −12.7 | +2.2 [0.7, 3.6] | 0.1 |
| 100 | 4 | +0.7 | +9.3 | +0.8 | +8.5 [6.5, 10.6] | 0.8 |
| 1,000 | 1.5 | −0.9 | −0.9 | −1.0 | +0.1 [−0.1, 0.4] | 0.1 |
| 1,000 | 2 | +0.6 | +0.7 | +0.0 | +0.7 [0.4, 1.0] | 0.3 |
| 1,000 | 4 | −0.9 | +6.1 | −1.2 | +7.2 [6.6, 7.9] | 2.4 |
| 10,000 | 1.5 | −0.3 | −0.2 | −0.1 | −0.1 [−0.2, 0.0] | −0.1 |
| 10,000 | 2 | −0.1 | +0.7 | −0.1 | +0.8 [0.7, 0.9] | 1.1 |
| 10,000 | 4 | −0.0 | +7.6 | −0.0 | +7.6 [7.4, 7.8] | 8.0 |
| *(b) Cut-off near the peak* | | | | | | |
| 100 | 1.5 | +0.1 | +3.0 | −0.2 | +3.2 [1.7, 4.7] | 0.1 |
| 100 | 2 | −12.5 | −12.3 | −14.0 | +1.7 [0.5, 2.9] | 0.0 |
| 100 | 4 | +0.9 | +10.1 | +4.8 | +5.3 [4.1, 6.5] | 0.2 |
| 1,000 | 1.5 | −1.4 | −2.0 | −2.1 | +0.1 [−0.2, 0.3] | 0.0 |
| 1,000 | 2 | −2.4 | −2.6 | −2.7 | +0.2 [0.0, 0.3] | 0.0 |
| 1,000 | 4 | −1.2 | +0.8 | −0.7 | +1.4 [1.1, 1.7] | 0.2 |
| 10,000 | 1.5 | −0.5 | −0.8 | −0.7 | −0.1 [−0.2, −0.1] | −0.1 |
| 10,000 | 2 | −0.1 | −0.4 | −0.3 | −0.1 [−0.1, 0.0] | 0.0 |
| 10,000 | 4 | −0.1 | +0.7 | −0.1 | +0.8 [0.7, 0.9] | 0.4 |

*simulated outbreaks*). Section S4.1 examines why it was below nominal.

#### S3.3 The case studies

For the three Ebola waves, the DDSA estimates of $R_0$ and of the removal rate were close to those of the continuous-time analysis of Vossler et al. (main manuscript, Section 3(d)). For H1N1 at WSU, the transmission rates agreed with those of KhudaBukhsh et al., but the population size and $R_0$ differed (main manuscript, Section 3(c)). The two analyses treated the population size differently: Algorithm 4.1 imputed $n$ at each iteration from the model-implied eventual infection probability, with a likelihood conditional on $n$, whereas DDSA summed $N$ out of the complete Binomial final-size term. To examine this choice, the WSU line list was fitted with the continuous-time re-implementation (exact times, with the 11 cases at $t = 0$ as initial infectives, whereas DDSA counted them as cases on the first day) under three population-size steps (Table S11). The imputation step of Algorithm 4.1 and the exact conditional draw under the complete Binomial final-size term shared the priors of the SDS example and the Metropolis-within-Gibbs sampler. The conditional draw was truncated to the support $N \leq 18{,}000$ of the DDSA analysis. With the imputation step, the estimates were close to those of the published code (Table S9). Replacing it with the exact conditional draw increased the estimated population size and lowered $R_0$, bringing both closer to DDSA, although this sampler mixed slowly (Table S11). Summing $N$ out gave similar estimates in the complementary comparison with NUTS and the priors of Section S3.2. These results suggest that the treatment of the population size accounts for much of the difference from the published analysis. The remaining difference from DDSA may reflect the daily records, the discrete map and the treatment of the cases at $t = 0$. This comparison did not separate their contributions.

Table S11: Population size and $R_0$ for the WSU H1N1 line list under three population-size steps with the same continuous-time likelihood (exact times, $T = 104$ d): posterior medians [95% CrI], with $N = n + m$. $\tau_\infty$: model-implied eventual infection probability of an initially susceptible individual at the current parameter values. $q_T$: probability of infection by $T$ for an initially susceptible individual. Last row: DDSA on daily records (main manuscript, Section 3(c)), posterior medians.

| Population-size step | $N$ | $R_0$ | Mean IP (d) | $\hat{R}$ |
|---|---|---|---|---|
| Imputation step of Algorithm 4.1, $n - K \sim \text{NegBin}(K, \tau_\infty)$, no $\binom{n}{K}$ | 7,786 [6,731, 9,052] | 1.09 [1.05, 1.14] | 5.13 | 1.008 |
| Exact conditional draw under the complete Binomial, $n - K \sim \text{NegBin}(K + 1, q_T)$ | 12,785 [9,391, 17,603] | 0.98 [0.94, 1.04] | 5.03 | 1.032 |
| $N$ summed out of the complete Binomial (NUTS) | 12,232 [9,092, 17,124] | 0.99 [0.94, 1.05] | 5.04 | 1.005 |
| DDSA (main text, Section 3(c)) | 14,144 | 0.97 | 5.12 | – |

##### S3.4 Practical advantages for daily line lists

Under the tested conditions, DDSA and continuous-time DSA gave similar estimates, except when the daily step was coarse relative to epidemic growth (Section S4.2). The advantages of DDSA for daily line lists are therefore mainly practical.

#### S4 Simulation checks of the stated limitations

The main manuscript attributes three limitations to specific components of DDSA: $R_0$ intervals that are narrower than nominal (the likelihood, L, shared with continuous-time DSA), a bias from the daily step when the epidemic grows quickly or the infectious period varies little (the map, M), and a cap on the number of stages of the GLCT map (G). This section reports the simulations behind each statement.

##### S4.1 Interval calibration

Outbreaks were simulated from a chain-binomial model that mirrors the GLCT map. New infections on day $k$ were $\text{Binomial}(S_k, 1 - e^{-\beta I_k/(n+1)})$, with $n$ initially susceptible individuals and one initial infective, and each case's infectious period consisted of $r = 4$ geometric stages with per-day progression probability $r/\overline{W}$ ($\overline{W} = 6.33$ d), which yielded the line list directly.

The grid was $n \in \{100, 1{,}000, 10{,}000\} \times R_0 \in \{1.3, 2, 3\}$ with 100 replicates per cell, conditioned on a major outbreak ($K \geq 0.1n$), and each replicate was fitted with the NegBin DPH model of the main manuscript and with the geometric model. The NegBin DPH model summed $r$ over $1, \ldots, \min(10, \min_i \delta_i)$. Each simulated stage lasted at least one day, so every recorded interval was at least 4 d and the true $r = 4$ was within the support. These fits used the $n$ initially susceptible individuals as the Binomial trials, with the whole-population infection probability $1 - \rho - S_T$, a slight variant of the convention of the main manuscript.

To examine the independence assumption of the likelihood, the NegBin DPH model was also fitted to outbreaks with the same parameters drawn from the DDSA working model, in which the number of cases followed the Binomial final-size term and the infection days and recovery intervals were drawn independently given the parameters. These outbreaks were not conditioned on a major outbreak, but every one had $K \geq 0.30n$.

Table S12: Calibration of DDSA with the GLCT map: $R_0$ (panel a) and mean infectious period (panel b) for the chain-binomial and working-model outbreaks of Section S4.1, fitted with the full line list. Bias: relative bias of the posterior median (%). Cov.: coverage of the 95% CrI. $k$: SD of the posterior medians across replicates divided by the mean posterior SD (95% interval width divided by 3.92), so $k \approx 1$ when the intervals match the variation between replicates. The coverage of the geometric model is in the repository's supplementary guide (guide, S4: *Baseline grid: all cells*). 100 replicates per row. An observed coverage of 0.95 has a 95% Wilson interval of about $[0.89, 0.98]$.

| | | Chain-binomial NegBin DPH | | | Working model NegBin DPH | | | Chain-binomial geometric |
|---|---|---|---|---|---|---|---|---|
| $n$ | True $R_0$ | Bias | Cov. | $k$ | Bias | Cov. | $k$ | Bias |
| *(a) $R_0$* | | | | | | | | |
| 100 | 1.3 | +1.4 | 0.38 | 3.0 | +0.3 | 0.94 | 1.1 | +7.5 |
| 100 | 2 | +1.2 | 0.59 | 2.4 | −0.1 | 0.94 | 1.1 | +14.7 |
| 100 | 3 | +0.9 | 0.80 | 1.6 | +1.6 | 0.97 | 0.9 | +20.4 |
| 1,000 | 1.3 | +1.1 | 0.28 | 6.0 | +0.1 | 0.90 | 1.0 | +5.2 |
| 1,000 | 2 | −0.5 | 0.51 | 2.7 | −0.0 | 0.94 | 1.0 | +11.4 |
| 1,000 | 3 | −0.1 | 0.67 | 2.0 | +0.5 | 0.97 | 1.0 | +17.7 |
| 10,000 | 1.3 | −0.2 | 0.17 | 6.9 | +0.0 | 0.95 | 1.0 | +4.6 |
| 10,000 | 2 | +0.4 | 0.52 | 2.9 | −0.0 | 0.96 | 1.0 | +12.2 |
| 10,000 | 3 | −0.2 | 0.68 | 1.9 | −0.0 | 0.95 | 1.0 | +16.9 |
| *(b) Mean infectious period* | | | | | | | | |
| 100 | 1.3 | −0.2 | 0.96 | 1.1 | −0.2 | 0.94 | 1.1 | −9.6 |
| 100 | 2 | −0.0 | 0.91 | 1.1 | −0.1 | 0.91 | 1.1 | −6.0 |
| 100 | 3 | +0.8 | 0.95 | 1.0 | −0.2 | 0.93 | 1.1 | −1.7 |
| 1,000 | 1.3 | −0.1 | 0.84 | 1.5 | −0.2 | 0.95 | 1.0 | −9.6 |
| 1,000 | 2 | −0.2 | 0.94 | 1.0 | +0.0 | 0.96 | 1.0 | −9.3 |
| 1,000 | 3 | +0.2 | 0.95 | 1.0 | +0.0 | 0.97 | 1.0 | −4.4 |
| 10,000 | 1.3 | +0.2 | 0.78 | 1.2 | +0.0 | 0.98 | 0.9 | −11.6 |
| 10,000 | 2 | −0.1 | 0.94 | 1.5 | +0.0 | 0.93 | 1.1 | −10.0 |
| 10,000 | 3 | −0.0 | 0.94 | 1.1 | +0.0 | 0.98 | 0.9 | −5.7 |

With the generating infectious-period family fitted, posterior medians were close to the truth, but nominal 95% intervals covered the true $R_0$ in only 17–80% of chain-binomial outbreaks, least often near the epidemic threshold (Table S12). Coverage on working-model data was much closer to nominal, with spread ratios near one.

DDSA and continuous-time DSA treat infection times as independent draws from the mean-field incidence, whereas in a stochastic epidemic later cases share the departures of the realised outbreak from that curve. The likelihood (L) does not represent this dependence. The comparison of the two generators supported, but did not isolate, its role, because the observation windows also differed, and the fitted final-size probability matched the working-model data but was $1 - \rho$ times the per-susceptible infection probability for the chain-binomial data. In the outbreaks of Section S3.2, a finer time step did not remove the under-coverage (Table S13), and in the chain-binomial outbreaks neither did a final-size term with extra-binomial variance (guide, S4: *Is the final-size term the cause of the narrow $R_0$ intervals?*).

Intervals for the mean infectious period were generally closer to nominal. In the larger chain-binomial outbreaks near the epidemic threshold, their coverage fell, whereas on working-model data it stayed close to nominal (Table S12b). The misspecified geometric model overestimated $R_0$ by 5–20%, more so at higher $R_0$, and underestimated the mean infectious period by 2–12% (Table S12).

#### S4.2 The daily step

Whether a daily step is adequate depends on the epidemic timescale relative to the step. Refining both the records and the map reduced the upward bias in $R_0$ in the fastest-growing setting, while estimates in the slower-growing settings changed little (Table S13). At $R_0 = 4$, $n = 10{,}000$ and the end cut-off, the difference between DDSA and continuous-time DSA was about eight posterior SDs, and no DDSA interval contained the true $R_0$ (Table S10). Refining the map alone would not recover timing information that daily records do not contain. These results point to the forward map step (M) rather than to the likelihood or the infectious-period structure.

The daily recording convention also raised the estimated mean infectious period by about 2%, because same-day recoveries were recorded as one-day intervals. This affected coverage most in the larger outbreaks, where intervals were narrow (guide, S3: *Mean infectious period*), and the difference from continuous-time DSA fell below 1% at $\Delta t = 0.25\,\mathrm{d}$ (Table S13b). With data ending at the peak, the fastest-growing setting ($R_0 = 4$) also underestimated the mean infectious period by about 12%, with intervals that rarely covered it. The step refinement was run only at the end cut-off, so whether this bias comes from the step was not tested.

Table S13: Convergence in the step $\Delta t$ of the discrete map: $R_0$ (panel a) and mean infectious period (panel b) from DDSA (geometric infectious period) with records and map at step $\Delta t$, for the outbreaks of Table S10 with $n = 1{,}000$ and the end cut-off. DDSA − DSA (exact): as in Table S10, as a percentage of the true value. It tends to zero as $\Delta t \to 0$, where the two likelihoods coincide. 50 replicates per row. Bias and coverage at every step are in the repository's supplementary guide (guide, S4: *Table S13: the daily step*).

| | DDSA − DSA (exact), % of truth [95% CI] | | | | Coverage of the 95% CrI | | |
|---|---|---|---|---|---|---|---|
| True $R_0$ | $\Delta t = 1$ | $\Delta t = 0.5$ | $\Delta t = 0.25$ | $\Delta t = 0.1$ | DDSA $\Delta t = 1$ | DDSA $\Delta t = 0.1$ | DSA (exact) |
| *(a) $R_0$* | | | | | | | |
| 1.5 | +0.1 [−0.1, 0.4] | +0.1 [−0.1, 0.4] | +0.1 [−0.1, 0.4] | +0.2 [−0.1, 0.4] | 0.36 | 0.36 | 0.42 |
| 2 | +0.7 [0.4, 1.0] | +0.3 [0.0, 0.6] | +0.1 [−0.2, 0.4] | +0.1 [−0.2, 0.3] | 0.42 | 0.46 | 0.52 |
| 4 | +7.2 [6.6, 7.9] | +3.4 [2.8, 4.0] | +1.7 [1.2, 2.3] | +0.8 [0.2, 1.3] | 0.54 | 0.80 | 0.84 |
| *(b) Mean infectious period* | | | | | | | |
| 1.5 | +2.8 [2.4, 3.1] | +1.1 [0.9, 1.3] | +0.5 [0.3, 0.7] | +0.2 [0.0, 0.4] | 0.84 | 0.86 | 0.82 |
| 2 | +2.1 [1.8, 2.4] | +0.6 [0.3, 0.8] | +0.1 [−0.1, 0.3] | −0.1 [−0.3, 0.1] | 0.74 | 0.84 | 0.84 |
| 4 | +1.4 [1.1, 1.7] | +0.2 [−0.1, 0.4] | −0.1 [−0.3, 0.1] | −0.1 [−0.4, 0.1] | 0.90 | 0.98 | 0.96 |

#### S4.3 The stage cap of the GLCT map

In the serial NegBin DPH of the GLCT map (G), an individual passes through at most one of its $r$ geometric stages per day, so the infectious period lasts at least $r$ days and the model requires $r \le \min_i \delta_i$. To see the effect of this cap on a low-variability infectious period, the synthetic setting of Di Lauro et al. (2022, §4.1) was used: $n = 9{,}950$ susceptibles and $m = 50$ initial infectives, $\beta = 0.25\,\mathrm{d}^{-1}$ and a Gamma infectious period with mean 9 d and variance 6 (shape 13.5), so $R_0 = 2.25$, observed to extinction, with 20 replicates. The outbreaks were simulated by the exact event-driven method of Section S3.2 and recorded by day in the same way. The data were the full line list (scenario A), infection times only with $N$ known (scenario B), or recovery intervals only (scenario C). Three models were fitted with the priors and sampler of Section S3.2.

The first was DDSA with the GLCT map, with $r$ summed out over $1, \ldots, \min(10, \min_i \delta_i)$. The second was DDSA with the Boxcar map and the discretised Gamma interval law implied by daily recording, whose support starts at one day for every shape. The third was the re-implemented continuous-time DSA with a Gamma infectious period.

Table S14: Effect of the minimum duration of the NegBin DPH law on DDSA estimates for the Gamma infectious period of Section S4.3 (mean 9 d, SD 2.45 d, shape 13.5, $R_0 = 2.25$), by scenario: full line list (A), infection times only with $N$ known (B) and recovery intervals only (C). Bias, Cov. and $k$ as in Table S12, with bias averaged over replicates. Gamma shape and $r$ ($r_{\max}$): medians over replicates of the posterior median shape, the number of stages in the GLCT model, and its upper bound (the shortest recorded interval). The number of stages and the Gamma shape are different parameters. 20 replicates per row, all fits included. In scenarios A and C, every fit had $\hat{R} \leq 1.01$, and at most 3 (A) and 1 (C) fits per model had divergent transitions. In scenario B, 6 Boxcar and 4 continuous-time fits had $\hat{R}$ of 1.01–1.02, and 5 and 10 had divergent transitions. An observed coverage of 0.95 has a 95% Wilson interval of about $[0.76, 0.99]$. $^{\dagger}$: not converged.

| | | $R_0$ | | | Mean IP | | | IP SD | | | |
|---|---|---|---|---|---|---|---|---|---|---|---|
| Scen. | Model | Bias | Cov. | $k$ | Bias | Cov. | $k$ | Bias | Cov. | Gamma shape | $r$ ($r_{\max}$) |
| A | DDSA, GLCT (NegBin DPH) | +10.4 | 0.00 | 5.5 | −3.0 | 0.05 | 3.9 | +104.3 | 0.00 | – | 2 (2) |
| A | DDSA, Boxcar (Gamma) | +3.8 | 0.00 | 2.7 | −0.3 | 0.85 | 1.1 | −0.2 | 0.90 | 13.5 | – |
| A | DSA (Gamma) | −0.3 | 0.40 | 2.7 | +0.0 | 0.95 | 1.1 | +0.3 | 0.90 | 13.5 | – |
| B | DDSA, GLCT (NegBin DPH)$^{\dagger}$ | *not converged (median $\hat{R} = 1.56$); not shown* | | | | | | | | | |
| B | DDSA, Boxcar (Gamma) | +0.2 | 0.85 | 1.3 | −16.5 | 0.05 | 1.8 | +19.8 | 0.70 | 6.5 | – |
| B | DSA (Gamma) | +0.3 | 0.85 | 1.3 | −0.1 | 0.75 | 1.6 | +5.5 | 0.85 | 14.8 | – |
| C | DDSA, GLCT (NegBin DPH) | – | – | – | −0.0 | 1.00 | 0.5 | +112.6 | 0.00 | – | 2 (2) |
| C | DDSA, Boxcar (Gamma) | – | – | – | −0.0 | 0.90 | 1.0 | +0.2 | 0.90 | 13.5 | – |
| C | DSA (Gamma) | – | – | – | −0.0 | 0.95 | 1.0 | +0.2 | 0.95 | 13.4 | – |

With the full line list (Scenario A), short recorded intervals restricted the GLCT fit to few stages. The fitted infectious-period distribution was too variable, and $R_0$ was overestimated with zero coverage (Table S14). The cap is a property of the NegBin DPH law as constructed here. A DPH obtained as $e^{A\Delta t}$ from a continuous-time phase-type distribution permits several transitions per step and does not impose the $r$-step minimum duration. Within the GLCT map, a mixture of serial branches with different numbers of stages could also accommodate a few short intervals without restricting the whole distribution. Neither was fitted here. Without the cap, NegBin DPHs with five or six stages would be close to the recorded Gamma law, but they give zero probability to about 2 and 6% of the recorded intervals, those shorter than five or six days (guide, S4: *The NegBin DPH law without the cap*). Larger samples increase the chance of recording short intervals. With recovery intervals only (Scenario C), the GLCT fit estimated the mean infectious period well but again overstated its variability, so the cap acted through the recovery term.

The Boxcar map (B) with the discretised Gamma law, fitted to the same daily data with the same likelihood (L) and map (M), has no such cap. It recovered the mean, variability and shape of the infectious period and reduced the bias in $R_0$, although the coverage of $R_0$ remained zero at the daily step (Table S14). Refining the records, map and interval law reduced the remaining bias and improved the coverage, which stayed well below nominal and similar to that of the re-implemented continuous-time DSA (guide, S4: *The remaining Boxcar $R_0$ bias and the time step*). This supported a discretisation explanation for the remaining bias, and suggested that a daily step can matter at moderate growth when the infectious period varies little. A Boxcar fit with the NegBin DPH law would share the minimum duration of the GLCT fit. The advantage shown here came from the Boxcar accepting an interval law whose support starts at one day, which in this setting was the generating family.

With infection times only (scenario B), the Boxcar fit estimated $R_0$ with little bias but underestimated the mean infectious period, with intervals that rarely covered it. In contrast, the re-implemented continuous-time DSA on exact infection times recovered it. The step refinement was run only for scenario A, so whether this bias comes from the daily records was not tested. The GLCT fits did not converge (Table S14).

### S5 Reproducibility and code

The Quarto notebooks for all analyses in the main manuscript and the code for the supplementary analyses are archived on Zenodo (`https://doi.org/10.5281/zenodo.23924832`). The vignettes of the Julia package *DiscreteDynamicSurvivalAnalysis.jl*, version 0.1.0 (`https://github.com/epirecipes/DiscreteDynamicSurvivalAnalysis.jl`), also reproduce the analyses of the main manuscript.

#### S5.1 Analyses

Each analysis was executed with Julia 1.11.6 and *Turing.jl* 0.49 in a single shared environment, whose package versions are recorded in the `Manifest.toml` at the root of the repository, together with two shared modules: `PhaseTypeDistributions.jl` (phase-type distributions) and `IntegerParameters.jl` (summing integer-valued parameters out of the likelihood).

DDSA models were sampled with NUTS (target acceptance 0.8, maximum tree depth 12, $\Delta_{\max} = 1000$, step size adapted by dual averaging). Each chain was started from jittered data-based values: the recovery parameter from the recorded intervals (the per-stage probability as $\min_i \delta_i/\overline{\delta}$ for the NegBin DPH and $1/\overline{\delta}$ for geometric models), the transmission rate from the final-size relation $R_0 = -\log(1 - K/N)/(K/N)$ where $N$ was known (and at $R_0 = 1$ where it was not), and the remaining parameters at their prior medians. In the Hagelloch analysis, the transmission rates started at their prior medians and the mean infectious period at the recorded mean. Every starting value was multiplied by $e^{0.1z}$, $z \sim \mathcal{N}(0, 1)$, independently per chain. The count-based comparators used four chains of 2,000 post-warm-up draws after 2,000 warm-up iterations, started from prior draws, with target acceptance 0.95 (0.8 with the removal rate fixed).

#### S5.2 Supplementary analyses

The analyses of Sections S2–S4 are in the folder `Supplementary_analyses/`. A guide notebook there, `supplementary_analyses.qmd` (rendered as `.html` and `.md`), states the design of each analysis, lists its scripts and the command that reruns them, and shows the tables and figures of this supplement. It also shows the analyses that this supplement summarises, under the headings cited in the text:

- S2: *Table S7: isolating the final-size term*; *Figure S1: the growth-rate ridge*; *The attack-rate sweep*; *Numerical grid posteriors: checks*; *Table S8 and Figure S2: larger populations.*
- S3: *Table S9: published codes in their own settings*; *Mean infectious period*; *Paired comparison and convergence.*
- S4: *Is the final-size term the cause of the narrow $R_0$ intervals?*; *Baseline grid: all cells*; *Table S13: the daily step*; *The NegBin DPH law without the cap*; *The remaining Boxcar $R_0$ bias and the time step.*
- S5: *Numerical validation and software.*

The script `run_method_supplement.sh` reruns these analyses in resumable chunks with fixed seeds and writes the tables of this supplement with `make_supplement_tables.py`.

The published code of KhudaBukhsh et al. and Di Lauro et al. is not included in the project repository. The script `S3_S6_dsa_comparison/get_authors_code.sh` downloads it at the commits that were run.

##### S5.3 Numerical validation and software

**Likelihood forms.** The DDSA fits of Sections S3 and S4 used a log-scale implementation that remained numerically stable in the far tails visited during warm-up. Near the posteriors of the main manuscript, it agreed with the implementation used there, and direct evaluations reproduced the factorisations of Eqs. (1) and (2).

**Grid posteriors.** The grids of Section S2 used unequal steps in the mean infectious period, finer at short periods. Their log densities were compared with those of the corresponding Turing models, and their sensitivity to grid refinement and to the support of the ODE's number of stages was checked (guide, S2: *Numerical grid posteriors: checks*).

**Continuous-time solvers.** The continuous-time mean field was integrated in log coordinates by fourth-order Runge–Kutta steps aligned with the infection times, and checked against an adaptive solver and the scheme of the published SDS code. A step-refinement example showed DDSA log-likelihood ratios approaching their continuous-time counterparts under the same final-size convention. The Gamma renewal scheme was checked under grid refinement and against the infection-time likelihood of the published Python code (Table S9).

**Software.** The published code was run unchanged, at commit `3ecb29d` of `SDS.Epidemic` (`https://github.com/cbskust/SDS.Epidemic`) and commit `26671d6` of `DSA_refactor` (`https://github.com/Zkeggia/DSA_refactor`), in the example settings of Section S3. Solver settings, check results, and the R and Python package versions are in the repository's supplementary guide (guide, S5: *Numerical validation and software*).
